# Biomarker-informed CSF proteomics reveals ENPP2–LPA lipid signaling associated with Alzheimer’s disease

**DOI:** 10.64898/2026.07.08.26357565

**Authors:** Min Qiao, Prabesh Bhattarai, Elanur Yilmaz, Alexander W. Rookyard, Lipi Das, Anu Jain, Dolly Reyes-Dumeyer, Annie J. Lee, Rafael Lantigua, Martin Medrano, Diones Rivera, Lawrence S. Honig, Lewis M. Brown, Caghan Kizil, Richard Mayeux, Badri N. Vardarajan

**Affiliations:** Taub Institute for Research on Alzheimer’s Disease and the Aging Brain; Department of Neurology; G.H. Sergievsky Center at the Vagelos College of Physicians and Surgeons, Columbia University, New York, NY; Department of Biological Sciences, Quantitative Proteomics and Metabolomics Center, Columbia University, New York, NY; School of Medicine, Pontificia Universidad Católica Madre y Maestra, Santiago, Dominican Republic; Department of Neurosurgery, CEDIMAT, Plaza de la Salud, Santo Domingo, Dominican Republic.; School of Medicine and Surgical Pathology, Universidad Pedro Henriquez Urena, Santo Domingo, Dominican Republic

## Abstract

**Background:** Alzheimer’s disease (AD) involves complex molecular alterations in the cerebrospinal fluid (CSF) proteome, yet the links between these protein changes and hallmark AD pathology remain incompletely defined. We investigated the relationship between the CSF proteome with CSF biomarkers of Alzheimer’s disease (AD).

**Methods:** CSF was collected in 500 individuals of non-Hispanic white, African Americans, and Caribbean Hispanic individuals. CSF biomarkers of AD were measured including P-tau181, Aβ40, Aβ42, total-tau, neurofilament light chain (NfL) and glial fibrillary acidic protein (GFAP). CSF was depleted of abundant proteins followed by precipitation, cysteine reduction/alkylation, and proteolytic cleavage by trypsin. Peptides were measured using a Q-Exactive HF mass spectrometer (Thermo Scientific). Association of individual and co-abundant modules of proteins were tested using evelated CSF P-tau181 and reduced Aβ42/Aβ40 to confirm the diagnosis of AD. We validated results in CSF from 397 participants in the Accelerated Medicine Partnership-Alzheimer’s Disease cohort. Associated proteins were functionally validated in postmortem human brains and zebrafish.

**Results:** We detected 1030 proteins, yielding an overall data completeness value of 97%. CSF levels of 75 (7.3%) proteins were significantly associated with CSF P-tau181 levels after multiple testing correction. Notably phospholipase D3 (PLD3, p=2.41E-09), apoE (p=4.25e-08) and osteopontin (OPN p=1.4E-16) were increased and autotaxin (ATX/ENPP2, p= 8.39E-09) and ceruloplasmin (CP) (p=2.72E-07) were lower among individuals with high P-tau181 levels. These proteins were also associated with CSF Aβ42/Aβ40 ratio and total tau levels but not with NfL. OPN was also associated with CSF levels of GFAP (p=1.32e-05).

Among proteins associated with P-tau181 levels, pathways related to axon development (p=2.4E-12), axonogenesis (p=1.45E-11) and regulation of axonogenesis (p=5.1E-09) were enriched. Immunostaining on postmortem human and zebrafish brain found that ENPP2 expression, the gene encoding ATX, was significantly reduced in AD brain and in the amyloidosis model in zebrafish. Reduced ENPP2 expression was consistent with reduced lysophosphatidic acid (LPA) levels in the CSF of individuals with AD. LPA administration into zebrafish CSF reduced the pathological changes in synapses and vasculature due to Aβ42.

**Conclusion:** Unbiased profiling of circulating CSF proteins among individuals with antemortem diagnosis of AD, identified key proteins PLD3, apoE, OPN, ATX, and ceruloplasmin. Validation in postmortem human brains and zebrafish models support potential roles in endosomal sorting and APP processing, inflammation, angiogenesis, lipid transport, and oxidative stress.

**One sentence summary:** Reduced autotaxin and lysophosphatidic acid were among 75 cerebrospinal fluid (CSF) proteins associated with biomarker-defined Alzheimer’s disease pathology.

## Introduction

Cerebrospinal fluid (CSF) biomarkers such as Aβ42, the Aβ42/Aβ40 ratio, phosphorylated tau at threonine 181 (P-tau181), neurofilament light chain (NFL), and glial fibrillary acidic protein (GFAP)^1–3^ are associated with Alzheimer’s disease (AD) pathology and amyloidosis on PET imaging. These biomarkers have become indispensable tools for AD diagnosis and monitoring pathophysiological changes *in vivo*. Despite their clinical utility, these core proteins capture only a subset of the molecular alterations occurring in AD^4^. Additionally, the majority of the proteomics studies in AD have been performed in brain tissue, which poses complexity due to the heterogeneity of cell types and regions and the agonal period prior to death.

Previous investigations of CSF proteomics have largely focused on identifying proteins differentially expressed between clinically diagnosed AD and controls, but this approach misses early disease changes that occur in preclinical AD before clinical symptoms emerge. By examining proteomic changes in relation to Core 1 AD biomarkers such as P-tau181 and amyloid-β levels, we capture pathological processes across the disease continuum and identify how these biomarkers are associated with other proteins related to pathogenesis. Additionally, few studies have functionally validated candidate proteins in experimental models to determine mechanistic relevance or potential therapeutic value.

We analyzed the relationship between untargeted CSF proteomics and established CSF biomarkers that define AD using validated CSF cut points^5^ and conducted functional experiments to provide mechanistic insights into pathways associated with antemortem AD pathology. Untargeted mass spectrometry-based proteomics was used to systematically characterize 897 CSF proteomes across four cohorts in relation to core CSF AD biomarkers including P-tau181, NFL, GFAP, and the Aβ42/Aβ40 ratio. We determined potential molecular pathways represented by biomarker-associated proteins that would nominate putative diagnostic and therapeutic targets in AD. Further, we conducted functional validation of AD biomarker-associated proteins in human brain and zebrafish models.

## Results

### Integrated CSF cohorts from diverse ancestries demonstrate consistent biomarker signatures of AD

CSF was collected in two cohorts- Caribbean Hispanics from the Estudio Familiar de la Influencia Genetica en Alzheimer (EFIGA) and non-Hispanic Whites from the CUMC Biobank who were evaluated for cognitive problems in the faculty practice at Columbia University. Participants were diagnosed using the NIA-AA^6^ criteria for clinical dementia attributed to Alzheimer’s disease. To enhance diagnostic specificity, we measured CSF P-tau181 and applied established cut points^5^ to identify participants with biomarker-positive AD and non-demented controls. Individuals that were demented and biomarker negative were deemed other dementias. We tested association of proteins with biomarker-positive AD, which provides greater biological certainty of underlying AD pathology.

This distinction is important because clinical dementia diagnoses, while based on standardized criteria, may include individuals with non-AD pathology that could influence the proteomic signature. 186 individuals were included from the EFIGA cohort of whom 59 (32%) were demented and 127 were cognitively unimpaired controls (Table 1). AD was present in 73 individuals and 222 individals with non-AD dementia were recruited from the CUMC CSF Biobank.

**Table 1:**
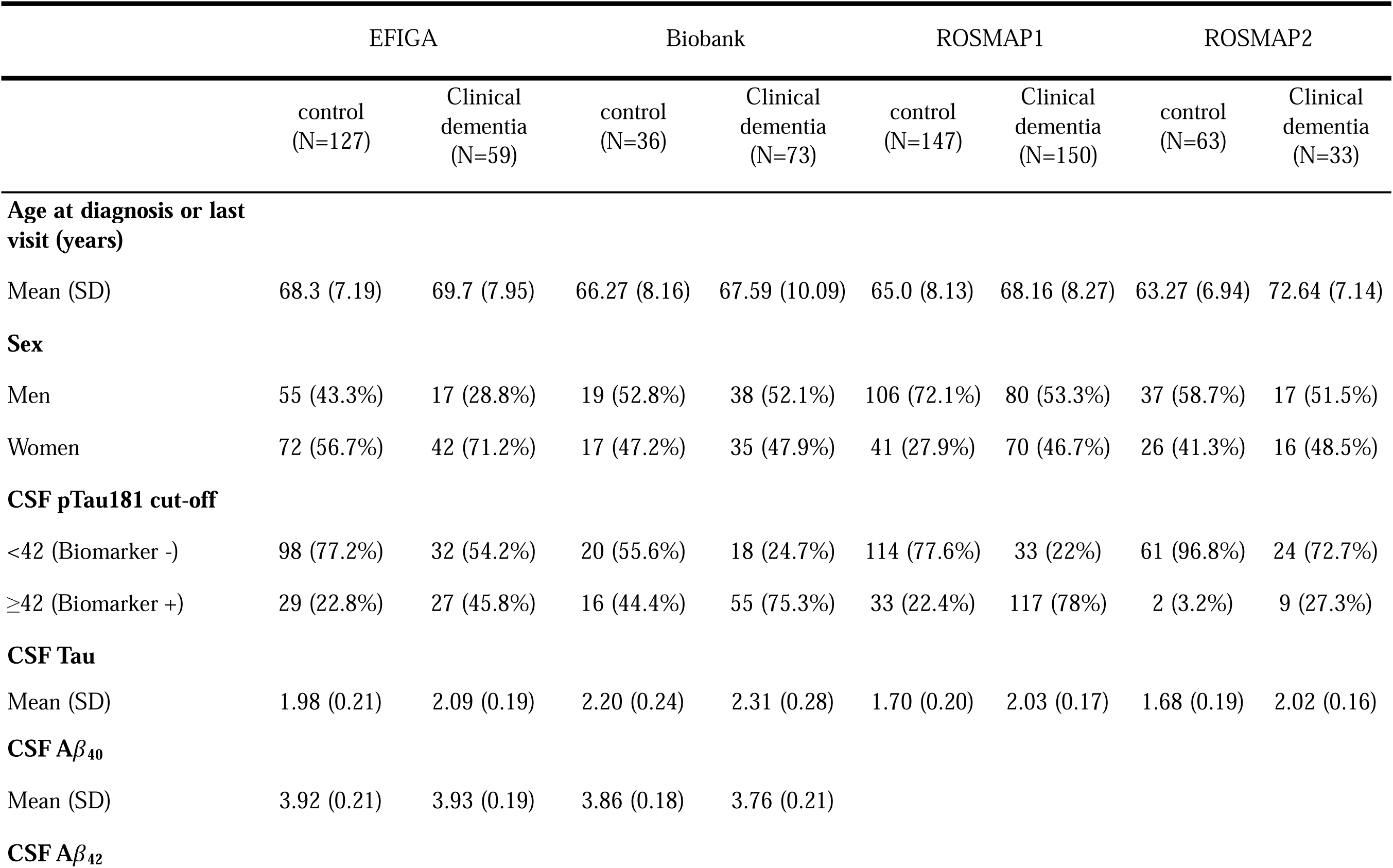
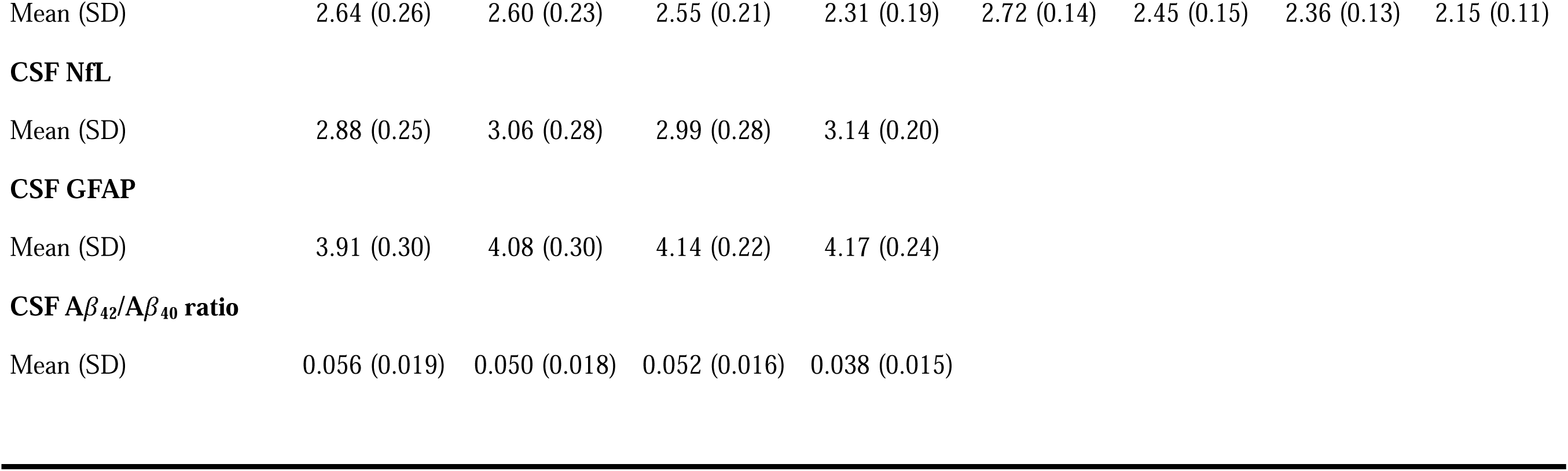
Demographics of participants included in the study.

|  | EFIGA |  | Biobank |  | ROSMAP1 |  | ROSMAP2 |  |
| --- | --- | --- | --- | --- | --- | --- | --- | --- |
|  | control<br>(N=127) | Clinical<br>dementia<br>(N=59) | control<br>(N=36) | Clinical<br>dementia<br>(N=73) | control<br>(N=147) | Clinical<br>dementia<br>(N=150) | control<br>(N=63) | Clinical<br>dementia<br>(N=33) |
| Age at diagnosis or last visit (years) |  |  |  |  |  |  |  |  |
| Mean (SD) | 68.3 (7.19) | 69.7 (7.95) | 66.27 (8.16) | 67.59 (10.09) | 65.0 (8.13) | 68.16 (8.27) | 63.27 (6.94) | 72.64 (7.14) |
| Sex |  |  |  |  |  |  |  |  |
| Men | 55 (43.3%) | 17 (28.8%) | 19 (52.8%) | 38 (52.1%) | 106 (72.1%) | 80 (53.3%) | 37 (58.7%) | 17 (51.5%) |
| Women | 72 (56.7%) | 42 (71.2%) | 17 (47.2%) | 35 (47.9%) | 41 (27.9%) | 70 (46.7%) | 26 (41.3%) | 16 (48.5%) |
| CSF pTau181 cut-off |  |  |  |  |  |  |  |  |
| <42 (Biomarker -) | 98 (77.2%) | 32 (54.2%) | 20 (55.6%) | 18 (24.7%) | 114 (77.6%) | 33 (22%) | 61 (96.8%) | 24 (72.7%) |
| ≥42 (Biomarker +) | 29 (22.8%) | 27 (45.8%) | 16 (44.4%) | 55 (75.3%) | 33 (22.4%) | 117 (78%) | 2 (3.2%) | 9 (27.3%) |
| CSF Tau |  |  |  |  |  |  |  |  |
| Mean (SD) | 1.98 (0.21) | 2.09 (0.19) | 2.20 (0.24) | 2.31 (0.28) | 1.70 (0.20) | 2.03 (0.17) | 1.68 (0.19) | 2.02 (0.16) |
| CSF Aβ<sub>40</sub> |  |  |  |  |  |  |  |  |
| Mean (SD) | 3.92 (0.21) | 3.93 (0.19) | 3.86 (0.18) | 3.76 (0.21) |  |  |  |  |
| CSF Aβ<sub>42</sub> |  |  |  |  |  |  |  |  |
| Mean (SD) | 2.64 (0.26) | 2.60 (0.23) | 2.55 (0.21) | 2.31 (0.19) | 2.72 (0.14) | 2.45 (0.15) | 2.36 (0.13) | 2.15 (0.11) |

**CSF NfL**
|  |  |  |  |  |
| --- | --- | --- | --- | --- |
| Mean (SD) | 2.88 (0.25) | 3.06 (0.28) | 2.99 (0.28) | 3.14 (0.20) |

**CSF GFAP**
|  |  |  |  |  |
| --- | --- | --- | --- | --- |
| Mean (SD) | 3.91 (0.30) | 4.08 (0.30) | 4.14 (0.22) | 4.17 (0.24) |

**CSF $A\beta_{42}/A\beta_{40}$ ratio**
|  |  |  |  |  |
| --- | --- | --- | --- | --- |
| Mean (SD) | 0.056 (0.019) | 0.050 (0.018) | 0.052 (0.016) | 0.038 (0.015) |

The study population had a mean age of 67.9 years (standard deviation (SD) = 8.4), the individuals with AD were slightly older, with a mean age of 68.6 (SD = 9.3), compared to controls who had a mean age of 67.9 (SD = 7.5). Women represented 51% of the cohort, and 45.2% of the cohort had one or more *APOE-*ε*4* alleles. Among patients diagnosed with dementia, 62% were biomarker positive using previously established cut points for CSF P-tau181for AD, while 28% of cognitively normal individuals that were biomarker positive. The mean levels of P-tau181, NfL and GFAP were higher in AD and the other dementias, than in controls while the ratio of Αβ42/Αβ40 was nominally lower (demographics and measurements in Table 1). There were significant differences in the CSF biomarkers (P-tau181, Aβ42/Aβ40 and Total-tau ratio) between patients with clinical diagnosis of AD and healthy controls in the CUMC Biobank and AMP-AD cohorts(Supplementary Figure S6). We also obtained postmortem brain proteomic data from a subset of participants from the ROSMAP cohort, 341 of whom were pathologically diagnosed with AD.

### Biomarker-anchored proteomic associations highlight AD-relevant pathways and potential therapeutic targets in AD

We detected and annotated 1030 proteins in the EFIGA and CUMC biobank cohorts. The two AMP-AD cohorts measured 431 of these same proteins. 26 (6%) proteins were associated with biomarker-positive AD (using CSF previously established P-tau181 cutoffs) (Figure 1A, Supplementary Table S1 and S5) in a meta-analysis of all four datasets. Notable associations include Autotaxin (ATX/ENPP2), Osteopontin (OPN), Apolipoprotein-E (apoE), ceruloplasmin (CP), Raf kinase inhibitory protein (RKIP), Phospholipase-D3 (PLD3) and lactate dehydrogenase B (LDHB). Results were largely unchanged when we repeated the association tests by excluding individuals that were biomarker negative other dementia, suggesting that the proteins were specific to antemortem AD pathology (Supplementary Table 6).

**Figure 1:**
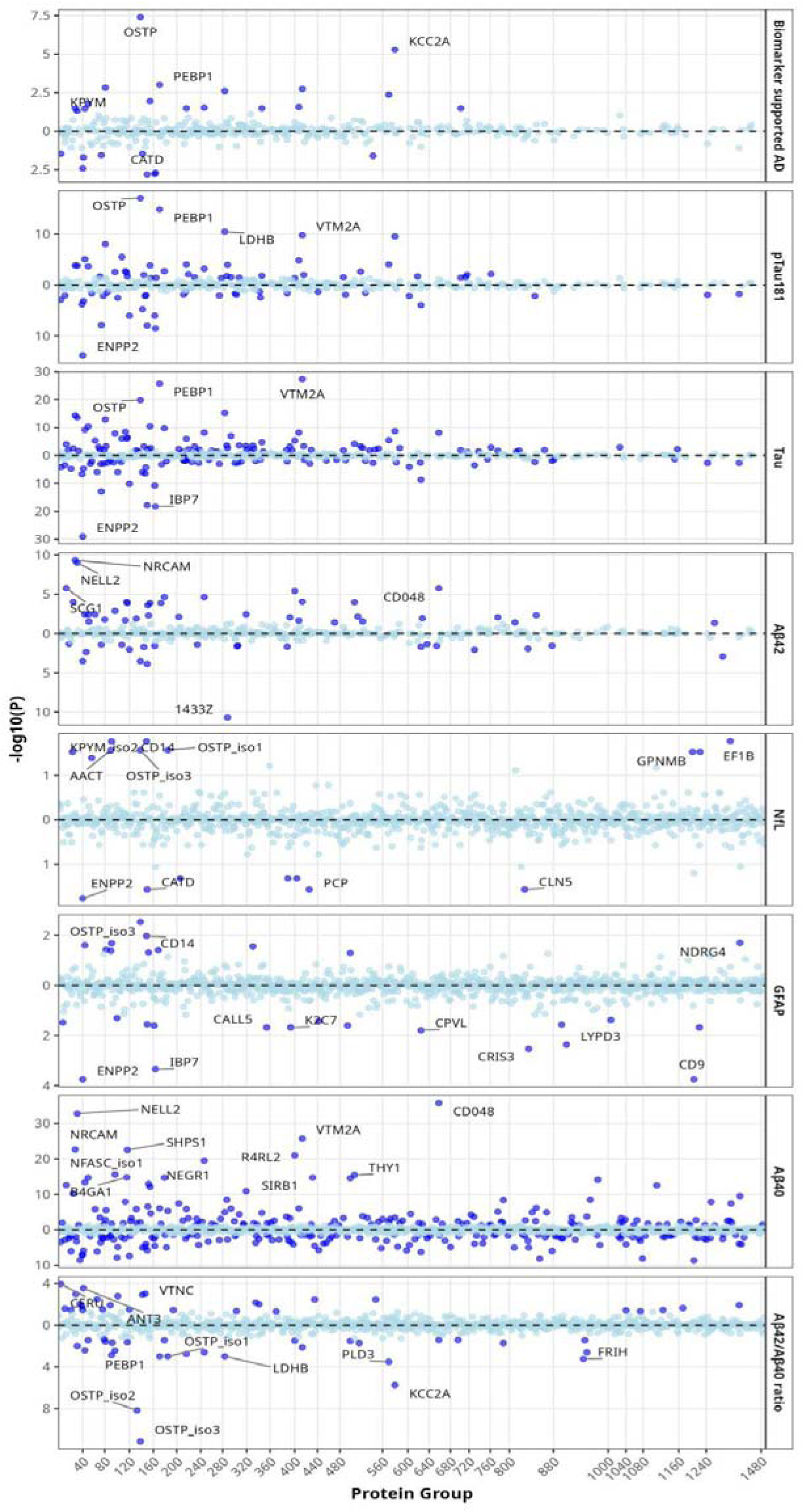
Manhattan plots of proteins (present in all datasets) and its association with clinical dementia, biomarker supported AD and all biomarkers (p-values are FDR corrected)

### Overlapping CSF protein networks connect P-tau181, T-tau, and amyloid biomarkers in AD

CSF P-tau181, total-tau (T-tau) and Aβ42 levels were available in all four datasets while we additionally measured CSF-NfL and CSF-GFAP in the EFIGA and CUMC Biobank cohorts. Among 431 proteins observed in all datasets, we observed statistically significant associations for 75 (17.4%) proteins with P-tau181 levels, 28 (6.5%) proteins with GFAP levels, 17 (3.9%) with NfL levels, 139 (32.3%) with T-tau and 56 (12.9%) with Aβ42/Aβ40 ratio (Figure 1A, Supplementary Figure S1, Supplementary Table 1). 25 proteins were associated with P-tau181, T-tau, Aβ42 levels and biomarker-positive AD (Figure 1B, Supplementary Table 1). Among cohorts where NfL and GFAP were also measured, we observed 19 proteins associated with P-tau181 and GFAP while only four proteins overlapped with P-tau181 and NfL (Figure 1B, Supplementary Table 1). OPN showed associations with all CSF AD biomarkers and biomarker-positive AD, whereas ENPP2 displayed selective associations with biomarker-positive AD indicating closer alignment with biological pathology than with clinical classification. LDH-B was associated with all phenotypes but not with Aβ42 and Aβ40.

**Figure 1B:**
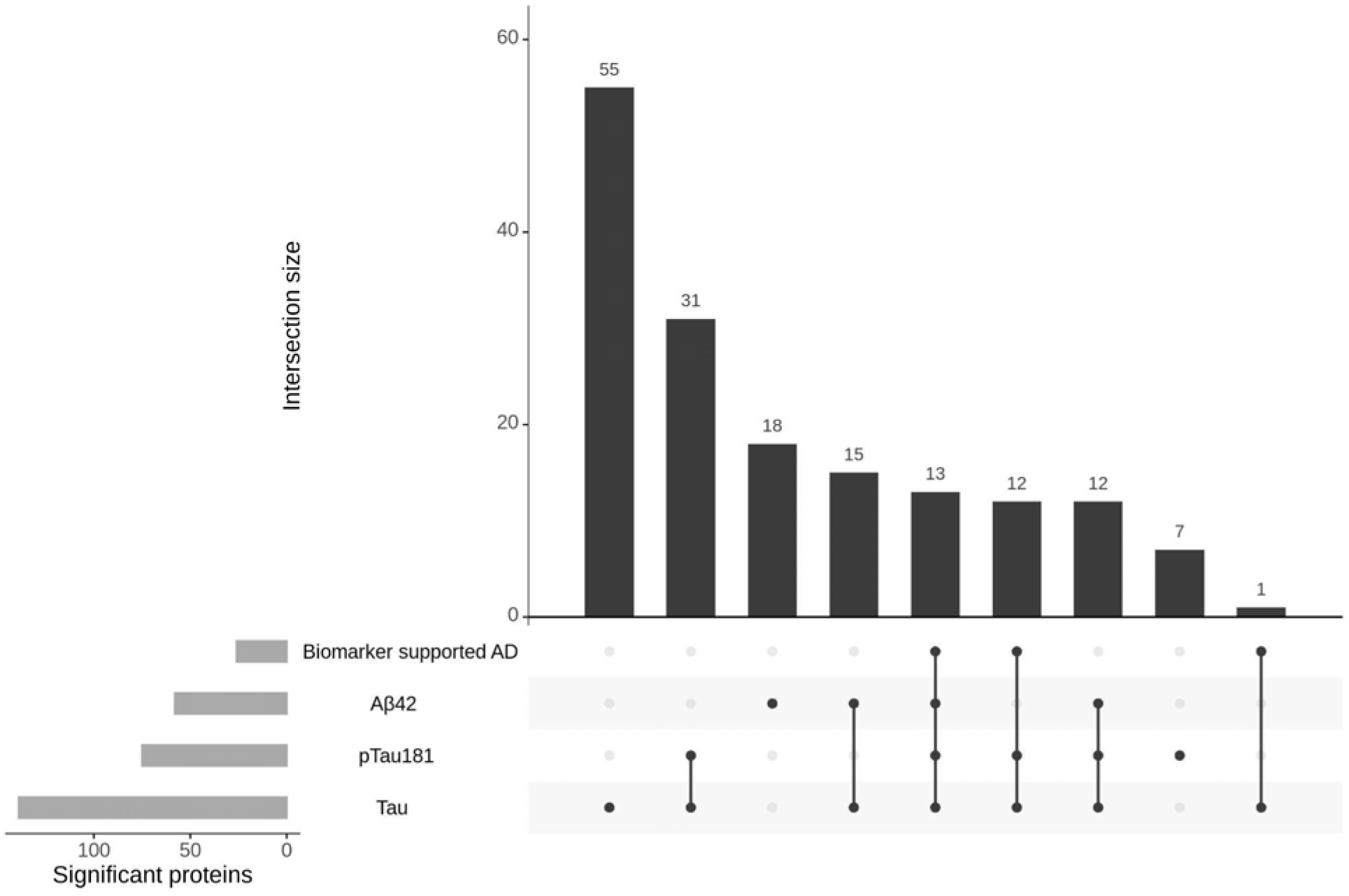
Upset plot of proteins associated with AD and CSF biomarkers across all datasets.

### Proteomic pathway analysis reveals vascular–inflammatory signatures and axonal remodeling in biomarker-positive AD

To investigate the biological relevance of our findings, we conducted pathway enrichment analysis of proteins associated with biomarker-positive AD and with CSF biomarker levels. We identified two distinct biological themes among proteins (Figure 2 and Supplementary Figure S2) associated with biomarker-positive AD, P-tau181, T-tau and other biomarkers of neurodegeneration. The first set of enriched pathways include cytoskeletal organization, synapse organization, axon development, and axonogenesis, indicative of alterations in neuronal structure, connectivity, and regenerative capacity. This reflects processes such as neurofilament release due to axonal damage or compensatory remodeling in response to injury^7–10^. A second biological theme included pathways related to wound healing, hemostasis, and blood coagulation. These pathways implicated vascular and inflammatory responses within the central nervous system. Such changes reflect disruption of the blood-brain barrier (BBB), extravasation of blood components, and the subsequent activation of immune, and clotting cascades that are processes observed in AD^8,11,12^, This distinct clustering of pathways suggests an interplay between vascular-inflammatory perturbations and disruption of neuron-specific structural or synaptic processes, which may collectively underlie neurodegeneration, impaired repair, or altered connectivity^8,9,12^.

**Figure 2:**
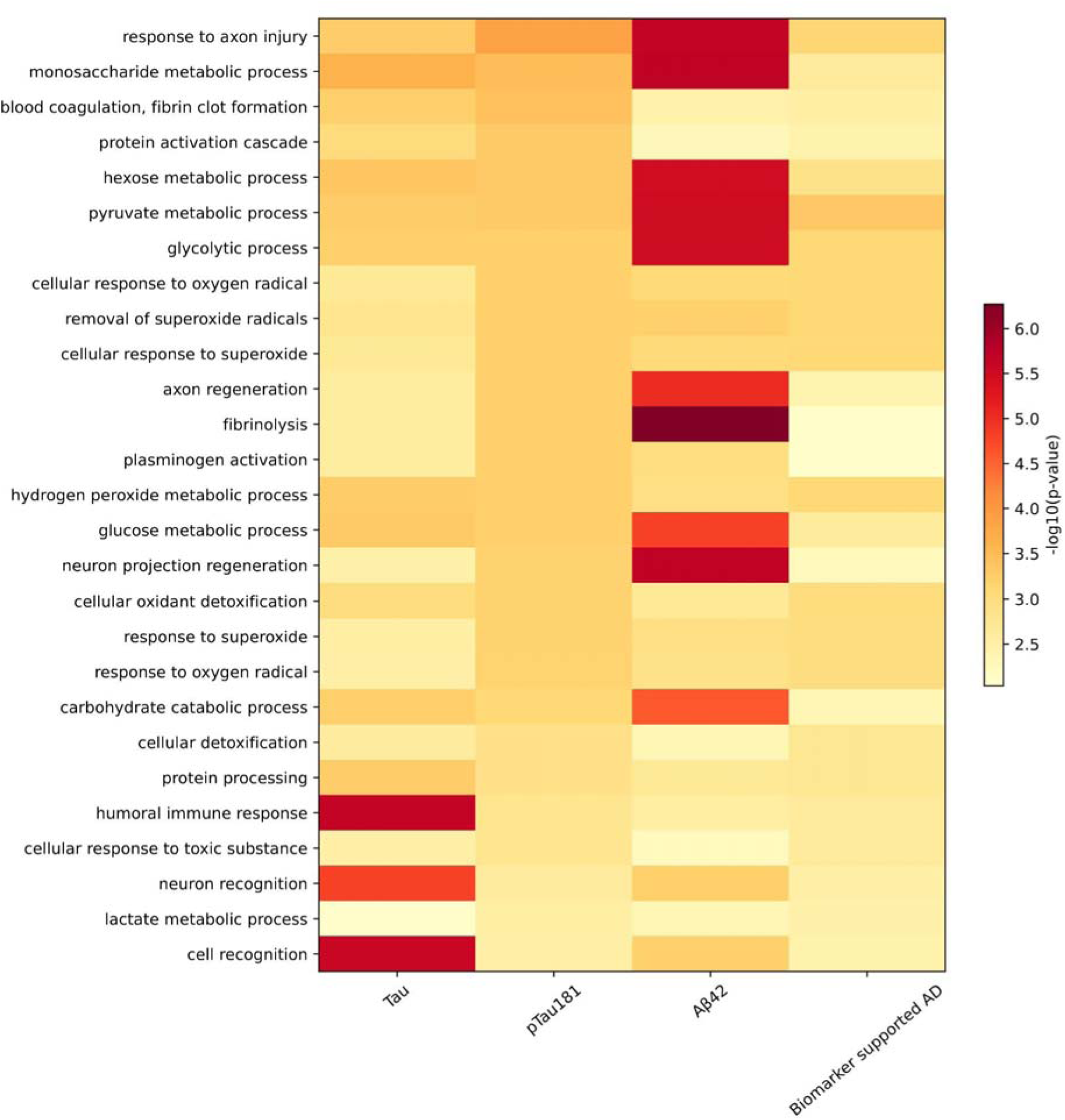
Pathways enriched amongst proteins significantly associated with biomarker assisted AD, P-Tau181, total-Tau and Aβ42 levels. Pathway enrichment was performed in 431 proteins that were characterized across all the datasets.

### Co-expression network analyses identified neuronal and vascular pathways associated with AD pathology

We clustered co-expressed proteins using weighted gene co-expression network analysis (WGCNA) to identify modules associated with CSF biomarkers and antemortem pathology. WGCNA identified 15 modules with at least 30 co-expressed proteins (Supplementary Figure S3, Supplementary Table S3). We then tested the association of each module with biomarker-positive AD and levels of the CSF biomarkers. The pink module which includes P-tau181 associated proteins-LHDB, PEBP1 and VTM2A, was positively associated with biomarker positive AD and all the CSF biomarkers. Additionally, salmon (PLD3) and midnight blue modules were also positively associated with biological AD, P-tau181, Aβ42/Aβ40 ratio and total-tau levels but not with NfL and GFAP. Green (CERU), greenyellow (OPN) and cyan (APOB) modules were negatively associated with biological AD, P-tau181, Aβ42/Aβ40 ratio and T-tau but not with NfL and GFAP. Pathway analyses of co-expression modules revealed that the positively associated pink, salmon and midnight blue modules contained proteins mostly enriched in neuronal pathways while negatively associated greenyellow, green and cyan modules contained proteins involved in vascular, extracellular and blood brain barrier related pathways (Supplementary Figure S4).

Using the top 25 most connected hub proteins in each module, we determined that the pink, salmon and midnight blue modules that are positively associated with biomarker supported AD and increased levels of CSF biomarkers were primarily expressed in excitatory and inhibitory neurons and oligodendrocyte precursor cells (Supplementary Table S4). The negatively associated green, greenyellow and cyan modules contained proteins expressed in astrocytes, oligodendrocytes, inhibitory neurons and oligodendrocyte precursor cells.

### LDH-B, PLD3 and APOE link CSF proteomic signatures to AD neuropathology

We validated CSF proteins associated with AD biomarkers by testing their association with brain pathology measured in the dorsolateral pre-frontal cortex from the ROSMAP postmortem collection. Several proteins associated with CSF biomarkers are also associated with post-mortem AD pathology (Supplementary Table S5, Supplementary Figure S5). LDH-B, PLD3 and apoE were associated with pathological AD and brain pathology (amyloid load and tangle burden), but the direction of effect was in the opposite direction compared to their effect in CSF. Interestingly, Heat Shock Protein Beta-1 (HSPB1) was strongly associated with global AD pathology, pathological diagnosis of AD, amyloid burden and tangle load were also associated with CSF levels of P-tau181. CERU was associated with CSF biomarker levels but was not associated with brain pathology.

### ENPP2 is selectively reduced in CSF, human brain, and amyloid-**β**–based zebrafish model

We accessed publicly available and in-house single-nucleus human and zebrafish brains to determine the expression profiles of *SPP1*, the gene that encodes OPN protein, and *ENPP2* in different brain cell types. *SPP1* is highly expressed in microglia and oligodendrocytes while *ENPP2* is predominantly expressed in oligodendrocytes (Supplementary Figure S7). Additionally, we also found that *ENPP2* expression levels were decreased in Aβ42 model of zebrafish, consistent with the findings in the CSF proteome.

We prioritized ENPP2 for functional validation in an amyloid-β–based zebrafish model^13–15^ because it emerged as the strongest association in our CSF proteomics analysis, showing significantly lower levels in biomarker supported AD compared to healthy controls. ENPP2 abundance was inversely associated with CSF P-tau181 contrary to evidence indicating that ENPP2 was increased in AD brain ^16^. Thus, we performed ENPP2 immunostaining on postmortem human brains (control, Braak stage 0 and AD, Braak stage V) and zebrafish brains (control versus Aβ42 injected) (Figure 3A). We selected zebrafish model due to its conserved neurovascular architecture, rapid *in vivo* imaging capabilities, and established utility in modeling AD-related pathology^14,15,17–31^. ENPP2 expression was significantly reduced in human brains with AD and zebrafish amyloidosis model (Figure 3 B).

**Figure 3:**
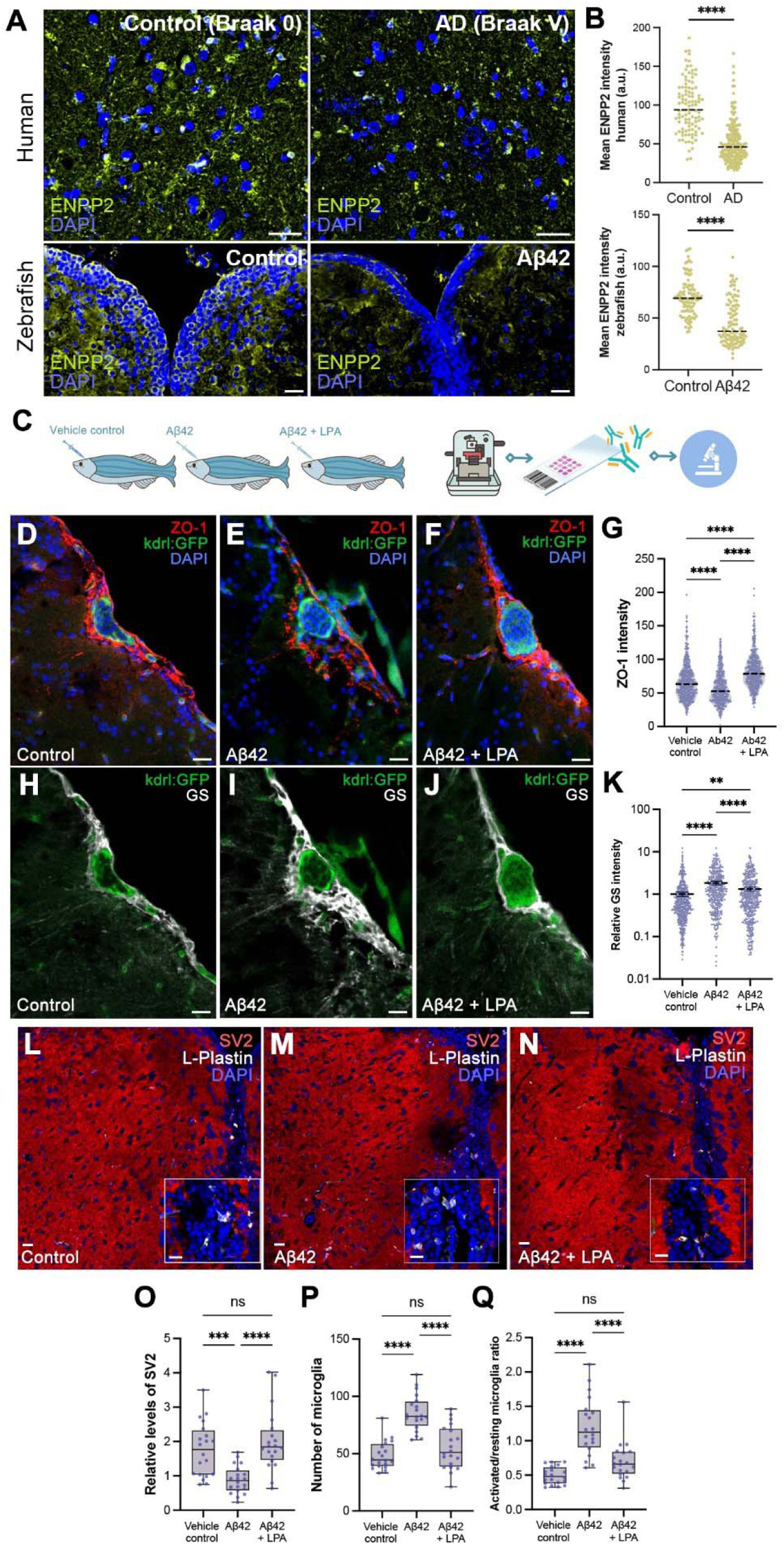
LPA ameliorates AD-related vascular, neural, and glial changes. (A) Immunostaining for ENPP2 with DAPI counterstain in postmortem human brains (control versus AD) and zebrafish brains (control vs Aβ42). (B) Quantification of mean ENPP2 intensity (Human: n = 4 brains, 104 cells in control, 187 cells in AD; Zebrafish: n = 8 brains, 99 cells in control, 112 cells in Aβ42). Mann-Whitney test, ****: p<0.0001. (C) Schematic overview of the functional analyses procedure in zebrafish. (D-F) Immunostaining for tight junction marker ZO-1 (red), and endothelial cell driven expression of GFP (kdrl:GFP, green) with DAPI nuclear counterstain in control, Aβ42-injected and Aβ42 + LPA injected zebrafish brains. (G) Quantification of ZO-1 intensities in endothelia. One-way ANOVA with Tukey post-test. ****: p<0.0001. (H-J) Immunostaining for astroglial marker GS (white), and GFP (kdrl:GFP, green) with DAPI nuclear counterstain in control, Aβ42-injected and Aβ42 + LPA injected zebrafish brains. (K) Quantification of GS intensity as a measure of reactive gliosis. One-way ANOVA with Tukey post-test. **: p<0.0021, ****: p<0.0001. (L-N) Immunostaining for synaptic marker SV2 (red), and microglial marker L-Plastin (white) with DAPI nuclear counterstain in control, Aβ42-injected and Aβ42 + LPA injected zebrafish brains. (O) Quantification of relative levels of SV2-positive synaptic puncta in L-N. (N) Quantification of the total number of microglia in L-N. (O) Quantification of the ratio of activated microglia to resting microglial as a measure of microglial activation. L-N: One-way ANOVA with Tukey post-test. **: p<0.0021, ***: p<0.0002, ****: p<0.0001; not significant (n.s.: p>0.0332). Scale bars equal 25 μm.

### LPA supplementation reverses amyloid-induced vascular, glial, and synaptic abnormalities associated with ENPP2

ENPP2 is the principal extracellular enzyme responsible for the generation of lysophosphatidic acid (LPA). Reduced LPA levels have been reported in the CSF of individuals with AD in independent studies^32^. In our analyses, ENPP2 levels were consistently reduced in biomarker-positive AD and in amyloid-exposed zebrafish (Figure 3A, B), raising the hypothesis that impaired ENPP2–LPA signaling contributes to AD-related pathology. To test the functional relevance of this pathway, we examined whether ectopic supplementation of LPA could mitigate amyloid-β–induced neuronal, vascular, and glial alterations *in vivo* (Figure 3C). We investigated how AD-related pathological changes, including reduced BBB integrity, elevated astrogliosis, increased microglial activation and more pronounced synaptic degeneration, change with LPA supplementation (Figure 3D-Q). We compared Aβ42-treated and LPA-injected animals to control and Aβ42-treated animals after performing immunostainings for vascular tight junction marker ZO-1, endothelial reporter kdrl-driven GFP, astroglial marker glutamine synthetase (GS), synaptic vesicle marker SV2 and microglial marker L-Plastin. We found that while Aβ42 reduced vascular integrity (lower ZO-1 expression in endothelia, Figure 3D-G), increased gliosis (higher GS expression levels, Figure 3H-K), hampered the synaptic density (lower number of SV2-expressing synaptic puncta, Figure 3L-O), and led to higher number of activated microglia (Figure 3L-N, P, Q), LPA treatment prevented the reduction in ZO-1 levels, reduced the increase in GS levels, prevented the reduction in SV2 positive synaptic puncta, and shifted the microglial activation state comparable to the control levels (Figure 3D-Q). These results suggest that reduced LPA because of amyloid toxicity could be among the prominent pathological culprits of vascular BBB dysfunction, astrogliosis, microglial activity and inflammation, and reduced synaptic density. Our results also validate the CSF proteomics findings in humans and imply that LPA supplementation could counteract AD related pathologies.

## Discussion

We identified proteomic alterations that track antemortem AD-related CSF biomarkers and facilitated prioritization of candidate pathways for mechanistic investigation. By extending biomarker-associated proteomic findings into an amyloid-β–based zebrafish model, we tested the functional relevance of a lipid signaling pathway implicated by human CSF data, while acknowledging the limitations of experimental models in fully recapitulating the complexity of human AD. Together, these findings provide a framework for linking human biomarker defined proteomic signatures to experimentally testable mechanisms relevant to AD-associated neurovascular and neuronal pathology.

Our proteome-wide analyses revealed several neuronal and synaptic pathways are associated with biomarker positive AD and with levels of specific biomarkers P-tau181, Aβ, total-Tau, NfL and GFAP. The association with biomarker-positive AD underscores the value of molecular phenotyping over conventional clinical categorization for detecting disease-specific alterations.

Pathway enrichment analyses delineated two complementary biological axes underlying AD pathology-neuronal and cytoskeletal processes, such as axonogenesis and synaptic organization, suggests neuronal remodeling or injury and vascular and inflammatory processes, implicating BBB disruption and immune activation. Vascular–inflammatory perturbation and neuronal structural dysfunction likely represent distinct but interacting molecular trajectories in AD. Targeting both vascular integrity and neuronal resilience may therefore offer synergistic therapeutic benefit across disease stages. Consistent with this, co-expression network analysis revealed neuronal modules positively associated with biomarker-positive AD and vascular–extracellular matrix modules inversely correlated with disease. These opposing associations suggest concurrent activation of neuronal stress responses and suppression of vascular-supportive processes. Such cell type specific patterns reinforce the concept that proteomic remodeling could occur even before overt symptoms, reflecting coordinated yet divergent cellular responses to neurodegeneration.

The consistent associations of OPN, ENPP2, and LDH-B across biomarker-positive AD highlight their potential as stage-specific biomarkers and therapeutic entry points. Although OPN showed broader associations with both biomarker positive AD and other dementia, its lack of specificity and established role as a general inflammatory marker led us to prioritize ENPP2, which demonstrated selective alignment with biological AD pathology and functional tractability. These proteins represent distinct molecular archetypes of AD and may inform biomarker panels tailored to disease stage and subtype.

Reduced ENPP2 levels in AD were validated in human and zebrafish brain tissue. Furthermore, we demonstrated that LPA treatment significantly mitigated Aβ42-induced vascular, neural, and glial changes associated with AD in the zebrafish model. LPA treatment preserved the integrity of the BBB, as evidenced by maintained levels of the tight junction protein ZO-1. This finding is crucial because vascular dysfunction contributes to AD by facilitating neuroinflammation and neuronal injury. Additionally, LPA reduced astrogliosis, as indicated by lower levels of the astroglial marker GS, and preserved synaptic density, as shown by the maintenance of SV2-positive synaptic puncta. Thus, LPA may not only protect against neuronal loss but also maintain the functional synaptic connections that are typically compromised in AD. Furthermore, the modulation of microglial activation states by LPA highlights its potential anti-inflammatory effects, which are vital in counteracting the chronic neuroinflammation observed in AD. Taken together, our findings suggest that augmenting LPA could serve as a therapeutic approach to address the complex pathology of AD. These in vivo findings provide functional validation of ENPP2’s role and support LPA as a potential candidate therapeutic target in AD. The next step would be to elucidate the precise molecular mechanisms underlying LPA’s protective effects and assess its therapeutic potential in clinical settings. These findings position ENPP2–LPA signaling within an emerging class of lipid-mediated pathways that may influence neurovascular integrity and synaptic resilience in AD.

The validation analysis using post-mortem brain tissue from the ROSMAP cohort revealed both convergent and divergent findings that highlight the complexity of CSF-brain protein relationships. While several CSF-associated proteins also associated with brain pathology, the opposite direction of effects suggested that CSF protein levels may not simply reflect brain tissue concentrations^33^.

Proteins measured in the brain may also be reduced in CSF by the neurodegenerative processes that decreases production and impaired transport from brain to CSF, or increased compartmentalization of the protein. Reduced levels often indicate neuronal loss or specific disease pathologies^34^, this discordance could indicate active secretion, differential clearance mechanisms, or the influence of BBB permeability on CSF protein composition. Future studies should validate these proteins longitudinally and develop targeted assays to integrate them into multimodal biomarker panels supporting precision medicine in AD.

## Methods

### Participants

The Estudio Familiar de la Influencia Genetica en Alzheimer (EFIGA) has been recruiting individuals with suspected sporadic and familial AD and healthy controls similar in age through advertisements in local newspapers and radio stations, and through clinical referrals in the Dominican Republic and in the Washington Heights neighborhood of New York City^5^. Participants in this study provided informed consent under protocols approved by the Columbia University Irving Medical Center Institutional Review Board, and the National Health Bioethics Committee of the Dominican Republic (CONABIOS). They underwent medical and neurological history and detailed examinations, neuropsychological testing, and collection of blood for plasma and DNA processing. CSF extraction was performed in a subgroup of participants participants both in the Dominican Republic and the Neurological Institute of New York. The clinical diagnosis of Alzheimer’s disease (AD) was based on NIA-AA criteria^35^. All clinical diagnoses were determined in a consensus conference attended by a neurologist, a neuropsychologist, and an internist with expertise in dementia and geriatrics. Briefly, individuals with clinical dementia must have a history of progressive cognitive decline in the absence of other brain disorders and objective evidence of a decline in memory and in at least two other cognitive domains such as verbal fluency or executive function. Healthy controls showed no evidence of cognitive decline. For the analyses in this manuscript, only biological samples and data from individuals recruited between January 1, 2018, and April 30, 2022, were considered.

### Blood and CSF collection in EFIGA and CUMC biobank

Blood for plasma was collected in dipotassium ethylenediaminetetraacetic acid tubes and centrifuged at 2000*g* for 15 minutes at 4 °C within 2 hours after collection. Plasma was aliquoted in polypropylene tubes, frozen, and stored at −80 °C. Blood for DNA extraction was also collected. Apolipoprotein E (*APOE*) genotyping was performed at LGC Genomics and CD Genomics. Cerebrospinal fluid was obtained with a standard aseptic technique, distributed into aliquots of 400 μL each in polypropylene tubes, frozen, and stored at −80 °C.

### Untargeted Proteomics

The CSF proteome was analyzed using mass spectrometry with data-independent acquisition and label-free quantification, providing a robust and scalable method to comprehensively characterize the proteomic landscape of the central nervous system. We describe the protocol for proteomic data generation and quality control in detail here^36^. All mass spectrometry raw data have been deposited in an international repository (MassIVE at https://massive.ucsd.edu).

### CSF based biomarker analyses

The methods have been previously described in detail^5^. Briefly, the CSF biomarkers assays were performed in duplicate using the SIMOA HD-X platform. Neurology 3-Plex A kits were used to determine levels of Aβ42, Aβ40, and T-tau, the Advantage V2 kit for P-tau181, and the Neurology 2-Plex B for GFAP and NfL. Ratio of Aβ42/Aβ40 was also calculated.

### Biomarker Positive/Biomarker supported AD

Based on previous laboratory-specific analysis^5^, P-tau-181 CSF level < 42pg/ml was considered biomarker negative (non-AD dementia or healthy controls) and ≥ 42 pg/ml considered biomarker-positive or biomarker-supported AD.

### Proteomics data processing

To address batch effects and batchwise variance in our proteomics dataset, we employed TAMPOR^37^ (Tool for Analyzing and Mitigating Proteomics Batch Effects and Outliers Robustly) for data correction and enable downstream analyses to focus on the biological variability of interest. The dataset consisted of 1,031 proteins measured across 500 samples. TAMPOR was applied with key parameters optimized for our study design: we utilized all non-GIS channels for correction (noGIS = TRUE, useAllNonGIS = TRUE) and specified median for central tendency adjustment. The algorithm iteratively adjusted the data over 250 iterations (iterations = 250), ensuring robust correction while maintaining the biological signal. To maximize computational efficiency, we utilized parallel processing across eight threads (parallelThreads = 8). A minimum batch size of five samples was set to avoid overfitting (minimumBatchSize = 5).

### Proteome-wide association study (PWAS)

PWAS was conducted using multiple linear models, adjusted for age and sex and first three principal components computed in the proteomics data. The analyses were conducted separately for proteins from each cohort, and we summarized the results in a random-effects meta-analysis using R library meta^38^. We report the p-value from the fixed effect ensuring there is no heterogeneity in the data. We corrected for multiple comparisons using an FDR of 5% and q-values were estimated using the Benjamini-Hochberg (BH) method.

### Proteome Co-abundance analysis: Co-abundance analysis of proteins

Co-abundance modular analysis was conducted using weighted gene correlation network analysis using the WGCNA R package^39^ (version 1.69). Using TAMPOR corrected abundance values for each proteomic feature from each sample, we first constructed a protein feature co-abundance network using pairwise Pearson correlations between each metabolic feature. We used a soft threshold of 4, chosen based on saturation of the R^2^ at 0.9. This correlation network, where the nodes are metabolic features and edges are the scaled correlation coefficients, was used to create the topological overlap matrix (TOM), which provides a measure of similarity between a given pair of metabolic features in the network. This similarity matrix was used to create a dendrogram to assign metabolic features into modules based on their co-abundance pattern. We used the following parameters: minimum module size of 10, merge cutHeight of 0.25, an unsigned network, and a reassign threshold of 0. After network and dendrogram construction, modules were defined using the *moduleEigengenes* function in WGCNA. The module eigengene is a quantitative representation of a module derived from a principal component analysis (PCA) as the first PC, conducted using only those metabolic features that were part of the module. Association analyses were conducted to find modules associated with outcomes in linear regression models, adjusted for age and sex. We used the Bonferroni method to correct for multiple comparisons.

### Hub proteins

To identify the top hub proteins for each protein module, we utilized the signedKME function from the R package WGCNA. This function calculates eigengene-based connectivity (kME) for all proteins, which reflects the correlation of each protein’s expression profile with the module eigengene (the first principal component of the module). Both positive and negative kME values were obtained, and we used their absolute values to prioritize top hub proteins, ensuring that both strong positive and strong negative correlations were considered. Our dataset consisted of 1031 proteins, grouped into 16 distinct protein modules, with each module containing at least 10 proteins. This approach allowed us to rank proteins within each module based on their centrality and functional relevance, facilitating the identification of representative hub proteins for downstream analysis.

### Pathway analysis

For pathway enrichment analysis, we employed the clusterProfiler^40^ package in R to identify significantly enriched biological processes, molecular functions, cellular components, and disease ontologies associated with differentially expressed proteins. Protein identifiers were converted to Entrez gene IDs using the biomaRt^41^ package. We performed separate enrichment analyses for upregulated proteins (fold change ≥ 1.5, adjusted p-value < 0.05) and downregulated proteins (fold change ≤ 0.67, adjusted p-value < 0.05). Gene Ontology (GO) enrichment analysis was conducted using the enrichGO function with the org.Hs.eg.db annotation database, applying Benjamini-Hochberg correction for multiple testing. Disease Ontology (DO) enrichment was performed using the enrichDO function. For visualization, we generated dotplots displaying the top 20 significantly enriched pathways for each category, where dot size represents the number of differentially expressed proteins in each pathway and color intensity indicates statistical significance. This approach allowed us to identify key biological processes and disease associations affected by our experimental conditions.

### CSF and brain proteomics processing in the ROSMAP datasets

For validation, we downloaded AMP-AD CSF proteomics data generated from two cohorts from Synapse (https://www.synapse.org/Synapse:syn21441787). We processed the proteomics data using the same criterion as mentioned above. We analyzed the AMP-AD two cohorts separately. Cohort 1 consisted of 297 samples with quantification of 528 proteins, while cohort 2 included 96 samples with 786 proteins. To capture the major axes of variation in protein abundance and enable downstream association analyses, we performed PCA independently within each cohort. The first three principal components (PCs) from each cohort were extracted and subsequently used as quantitative traits in association testing with Alzheimer’s disease biomarkers (Aβ42, total-tau, and P-tau 181), as well as clinical Alzheimer’s disease status.

In addition to CSF-based proteomics, we analyzed brain proteomics data generated from the dorsolateral prefrontal cortex (DLPFC) in the ROSMAP cohort^42^ (https://www.synapse.org/Synapse:syn21449447). This dataset comprised 8,425 proteins quantified across 563 postmortem brain samples. The brain proteome data were primarily used to evaluate and replicate protein associations identified in CSF. As part of the neuropathological characterization, each sample was accompanied by measures of amyloid plaque load and tangle burden, which were used as outcome traits in association analyses with protein abundance. Protein expression data were log-transformed and adjusted for covariates as needed, and association analyses were performed in parallel with those conducted in CSF.

### Modeling ENPP2 in Zebrafish

#### Housing and maintenance of animals

All animal experiments were conducted in compliance with applicable regulations and were approved by the Institutional Animal Care and Use Committee (IACUC) at Columbia University and/or the Norwegian Food Safety Authority. Columbia University holds the following accreditations: Columbia University Assurance - #D16-00003 (A3007-01), Columbia University USDA Registration - #21-R-0082, Columbia University AAALAC Accreditation - #000687, and Columbia University NYDOH - #A141. The animal care and use program at Columbia University is accredited by AAALAC International and maintains an Animal Welfare Assurance with the Public Health Service (PHS), Assurance number D16-00003 (A3007-01). Animals were handled with caution to minimize suffering and reduce the overall number of animals used.

#### Randomization

To minimize selection bias, zebrafish were randomly assigned to experimental and control groups, ensuring equal chances for placement in any group and comparability at the experiment’s start. In experiments with known or potential confounding variables (e.g., age, sex, or batch effects in transgenic lines), a randomized block design was utilized. Zebrafish were grouped into blocks based on known variables, and individuals within each block were then randomly assigned to experimental groups.

#### Health status

The full-time dedicated veterinary staff, trained and experienced with the species used in the University research programs, are available 24/7. Animals are checked daily, with more frequent monitoring as needed. An intensive care unit provides continuous monitoring and medical care if necessary. Within the University program, the Attending Veterinarian or their delegates are authorized by the Institutional Official to intervene to alleviate pain or distress as deemed appropriate. The health status of animals was regularly checked observationally, pathologically, and molecularly, with no health issues detected for the animals used in this study.

#### Treatment and tissue preparations

Cerebroventricular microinjections (CVMI) were conducted on adult zebrafish brains, following established protocols^15,22,24,28^. Endothelial kdrl transgenic reporter zebrafish (Tg(kdrl:GFP)^43^, aged 9 months and of both sexes, were used in the experiments. The animals were injected with PBS (control), human amyloid-beta42 (Aβ42, 20 µM), or LPA (10 μM), with a total injection volume of 0.5 – 1 μl. Three days post-injection (dpi), the fish were euthanized and their heads were dissected and fixed overnight at 4°C in 4% paraformaldehyde. After several washes, the heads were incubated overnight at 4°C in a solution of 20% sucrose and 20% ethylenediaminetetraacetic acid (EDTA) for cryoprotection and decalcification. The following day, the heads were embedded in O.C.T. compound and cryosectioned into 12-μm thick sections on SuperFrost Plus glass slides. For immunohistochemistry, sections were dried at room temperature, washed with PBS containing 0.03% Triton X-100 (PBSTx), and incubated overnight at 4°C with primary antibodies. The next day, the slides were washed with PBSTx and incubated with secondary antibodies for 2 hours at room temperature, followed by multiple washes before mounting with 70% glycerol in PBS.

#### Imaging and quantifications

Images were captured using a Zeiss LSM800 confocal microscope at 20X magnification with tile/z-stack functions as necessary. For quantitative analyses, five telencephalon sections between the caudal end of the olfactory bulb and anterior commissure were used per animal (n=4 animals per experimental group). Synaptic vesicle protein 2 (SV2)-positive synapses were quantified using the 3D object counter module in ImageJ software, applying a uniform threshold across all images. Microglia were quantified by counting L-Plastin immunoreactive cells. For glial marker Glutamine synthetase (GS) and zonula occludens-1 (ZO-1), regions of interest (ROI) were identified using GFP labeling in Zeiss ZEN software, and fluorescence intensities for specific channels were exported as data tables for each ROI (>400 ROIs analyzed per group) as described before^17–19,44–46^. Image acquisition was performed in a blinded manner. Statistical analyses were conducted using GraphPad Prism (Version 9.5.1) with one-way ANOVA and appropriate post-tests based on data structure (Sidak’s, Tukey’s, or Dunnett’s multiple comparison tests, Brown-Forsythe and Welch post-test, or non-parametric Kolmogorov-Smirnov test). Error bars represent s.e.m., with significance indicated as follows: *: p<0.0332, **: p<0.0021, ***: p<0.0002, ****: p<0.0001, and not significant (n.s.: p>0.0332).

## Data Availability

All data produced in the present study are available upon reasonable request to the authors

https://www.synapse.org/Synapse:syn21441787

https://www.synapse.org/Synapse:syn21449447

https://cumc.co1.qualtrics.com/jfe/form/SV_dmck0uV3A91pmzb

