## Supplementary Figures S1-S7 for "Biomarker-informed CSF proteomics reveals ENPP2–LPA lipid signaling associated with Alzheimer’s disease"

**Figure S1: Overlap between proteins significantly associated with AD and AD biomarkers measured in the EFIGA and CU Biobank cohorts**

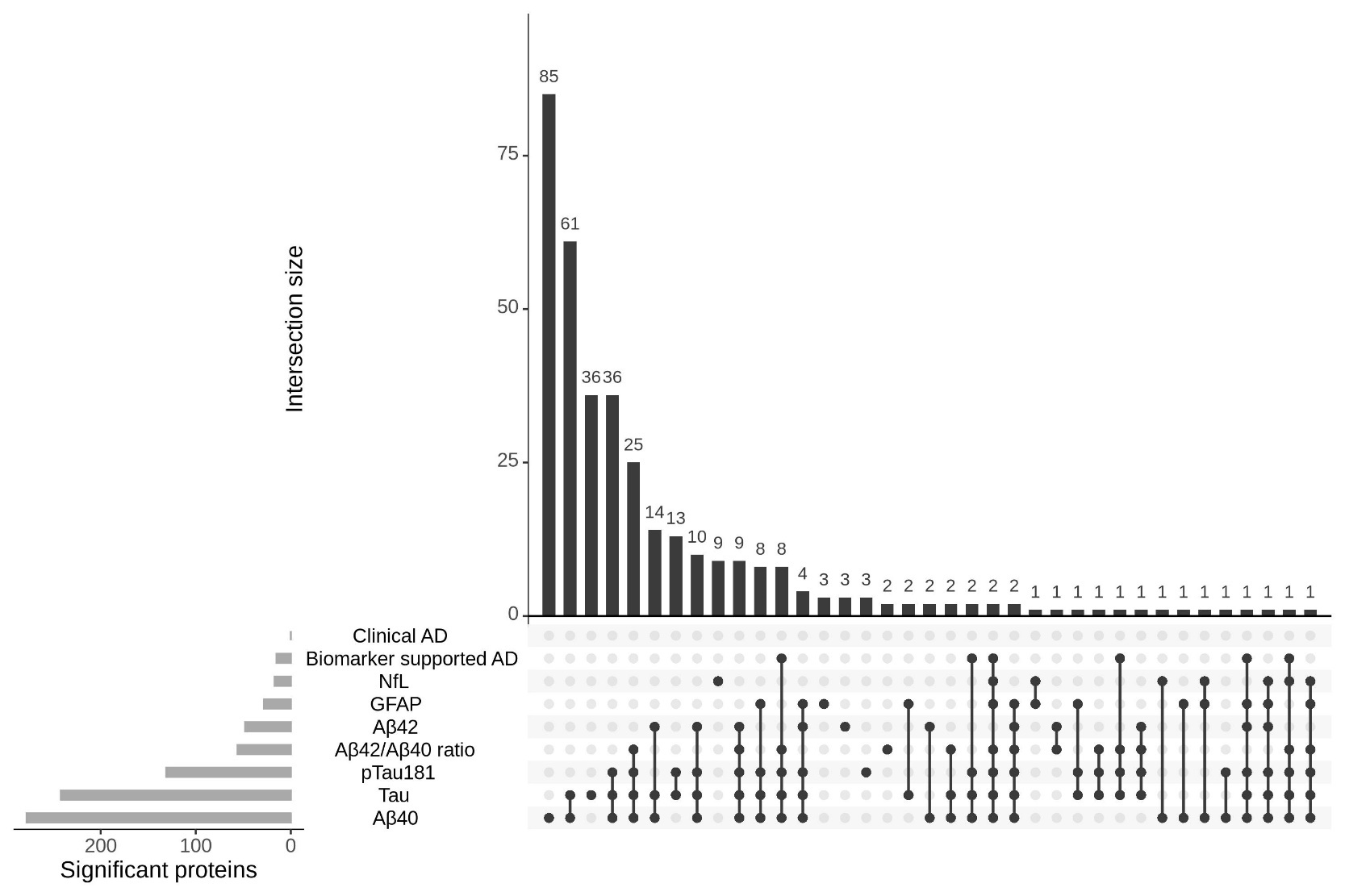

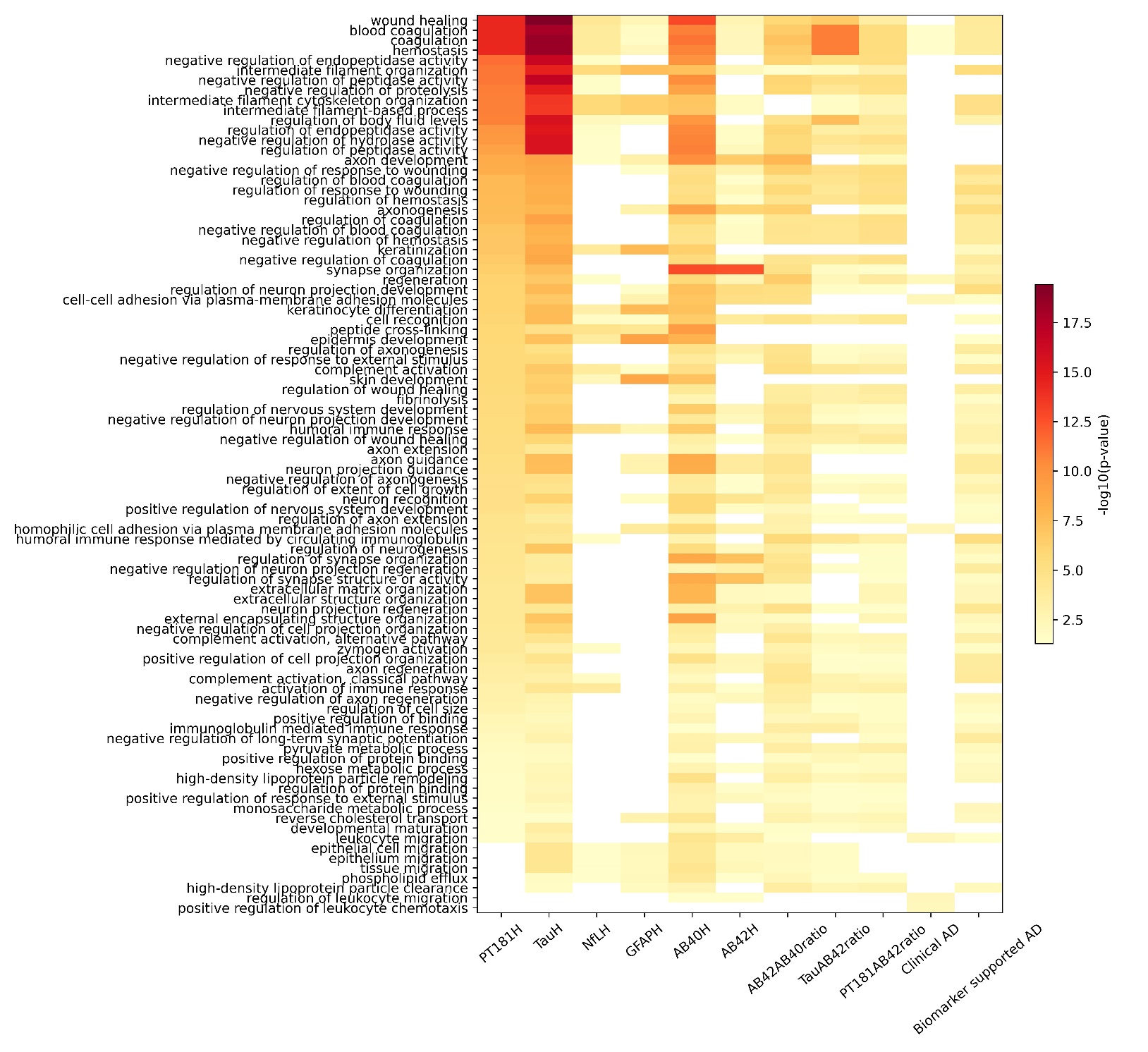
**Figure S2: Pathways enriched amongst proteins associated with AD and biomarkers**

**Figure S3: Modules associated with CSF biomarkers, clinical and biomarker supported AD. Panel B shows levels the top hub proteins in significantly associated modules in Biomarker positive and negative participants.**

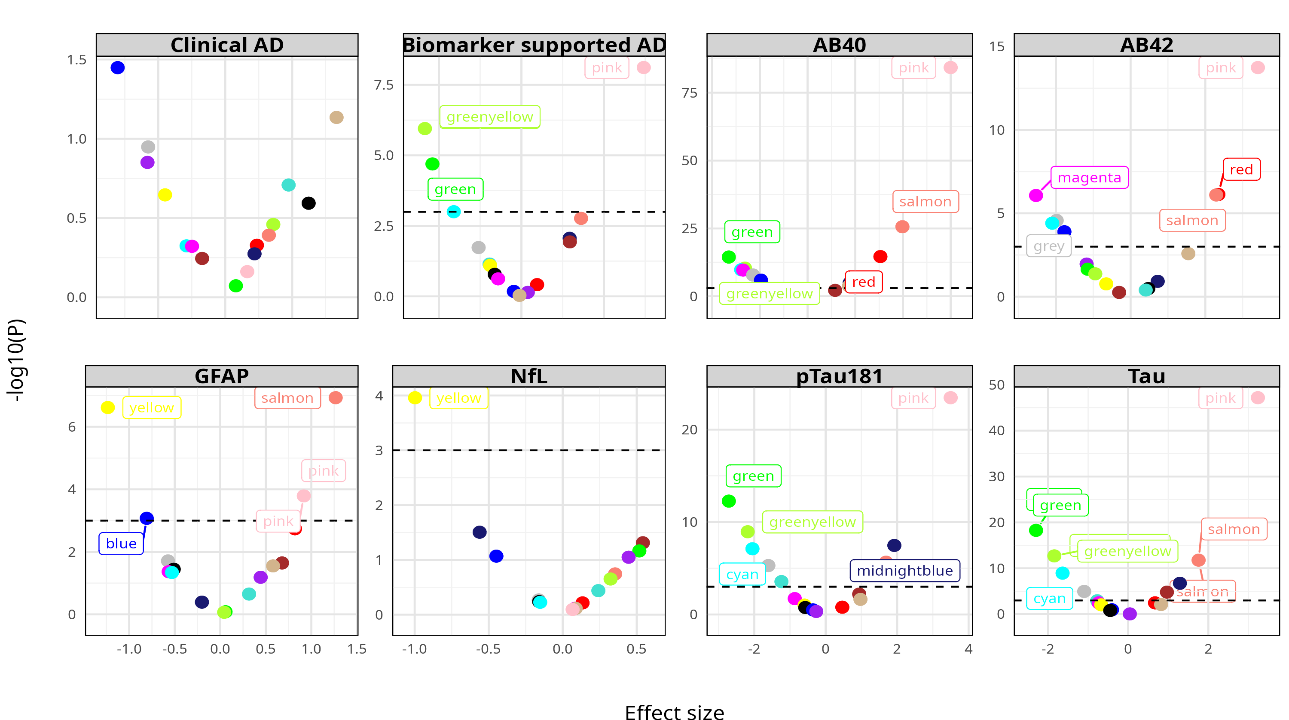

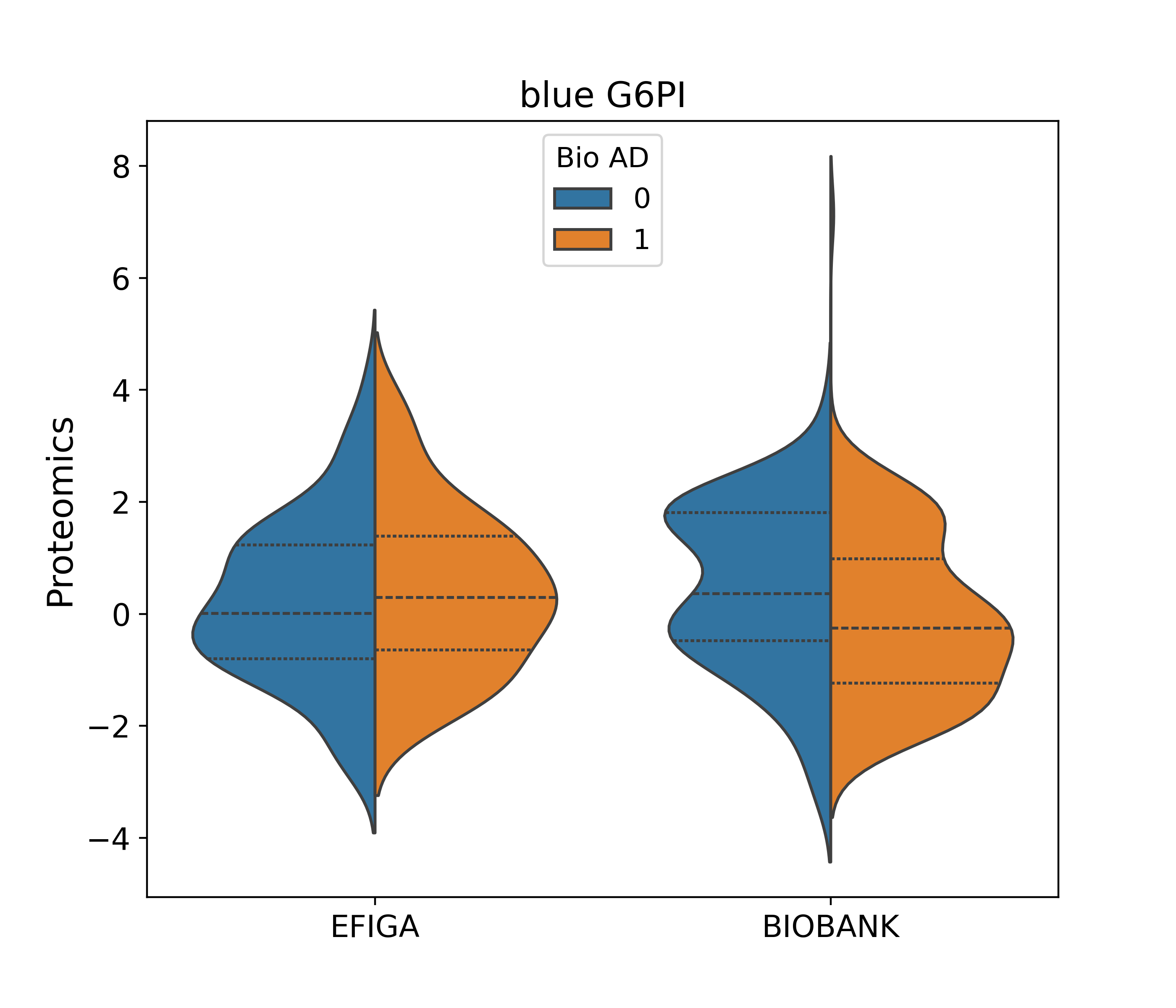

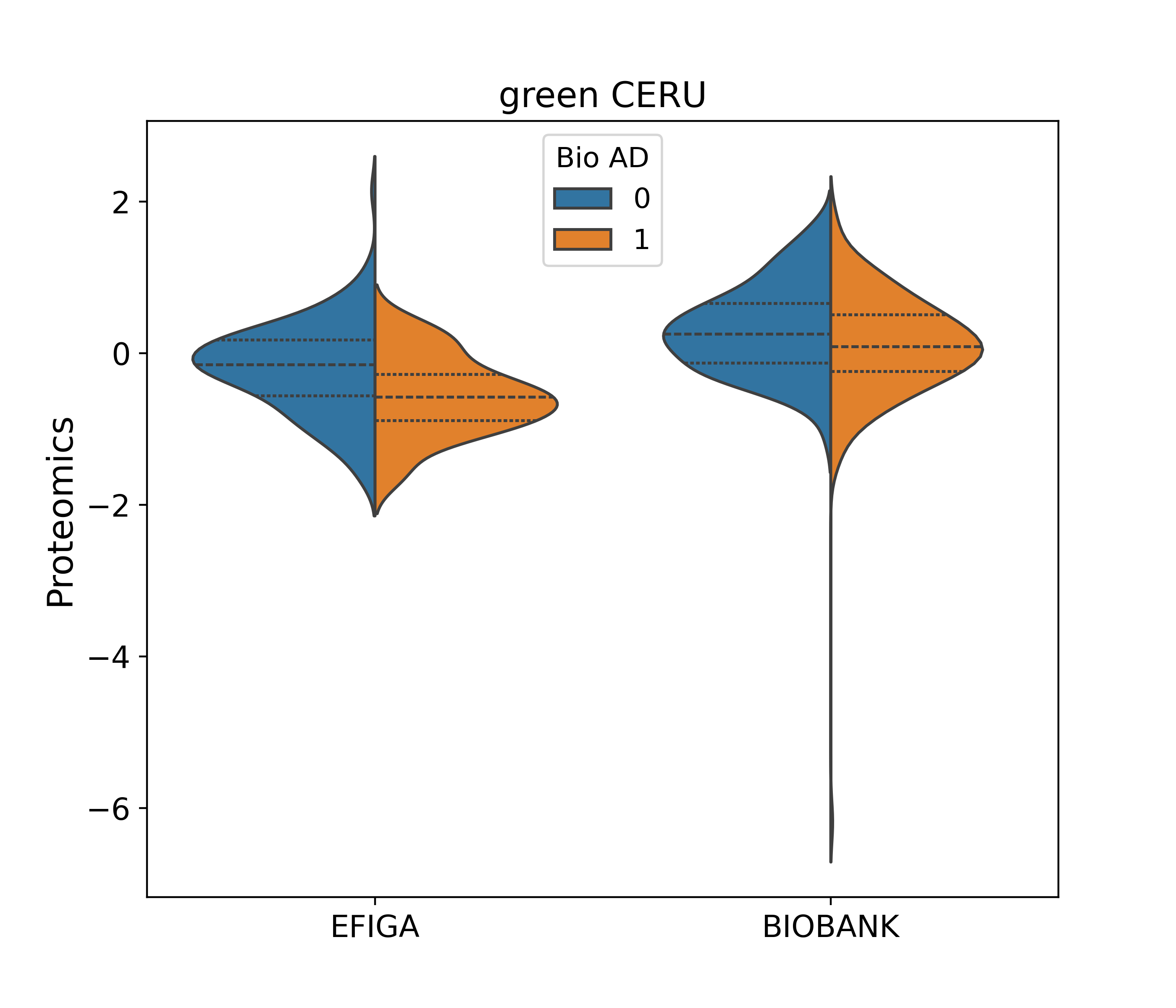

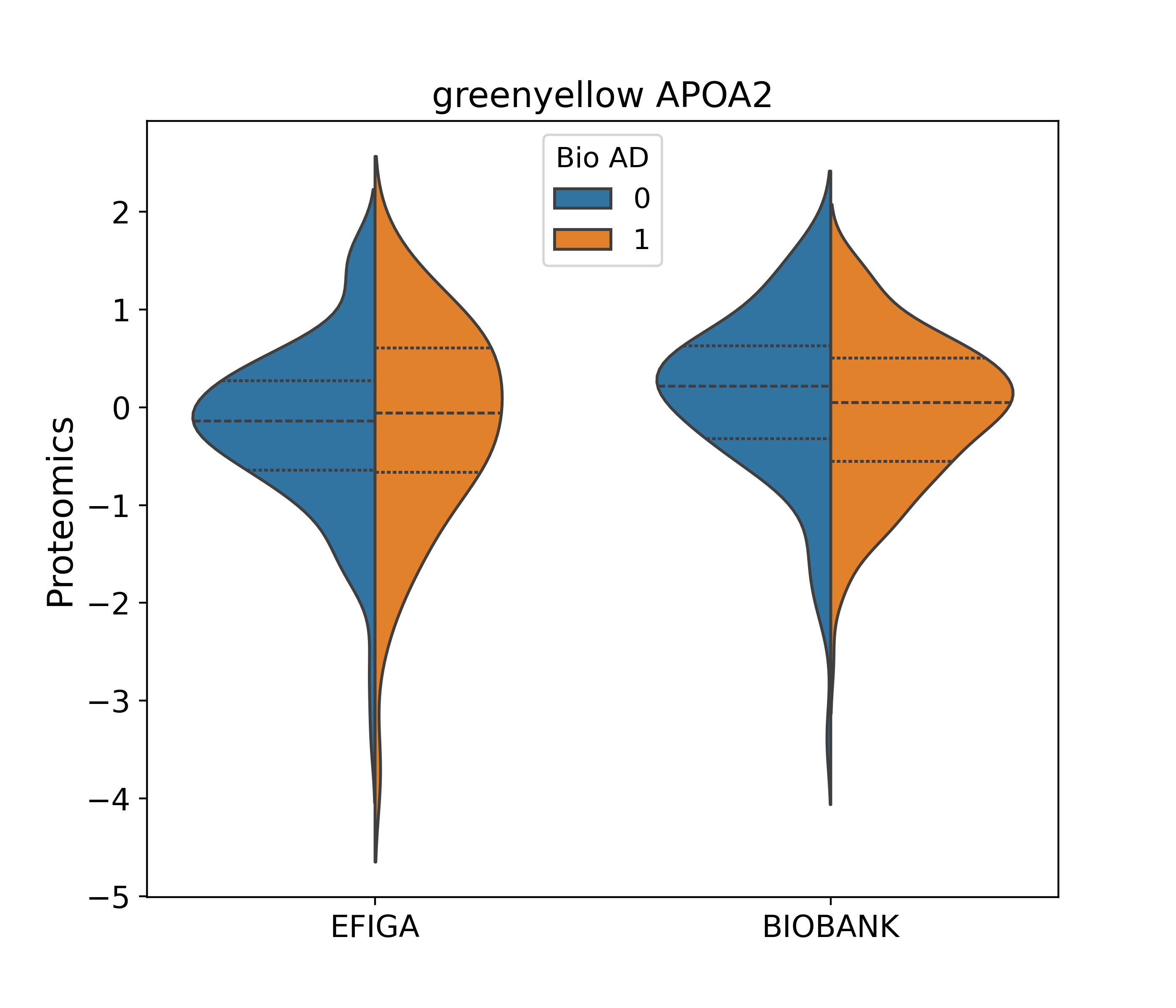

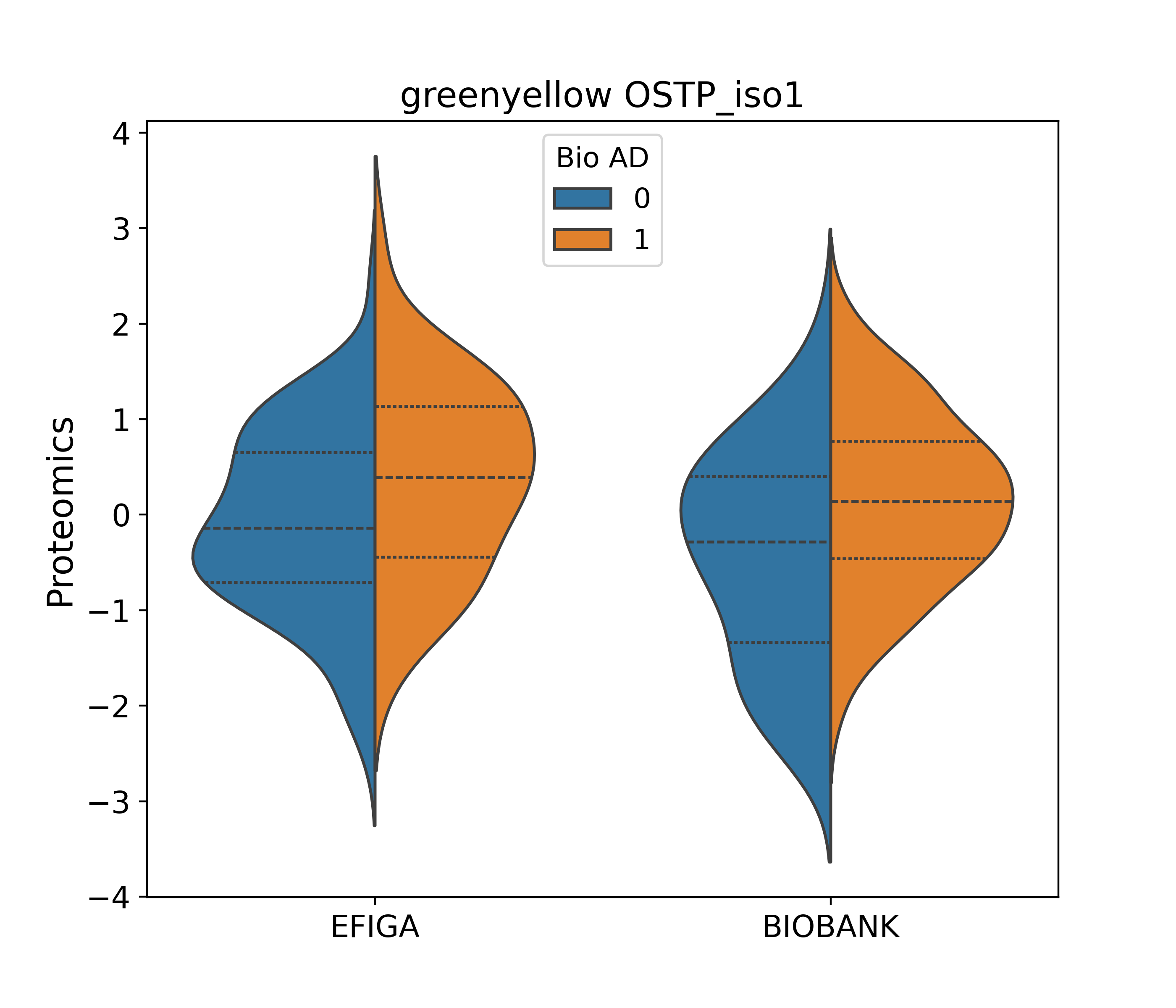

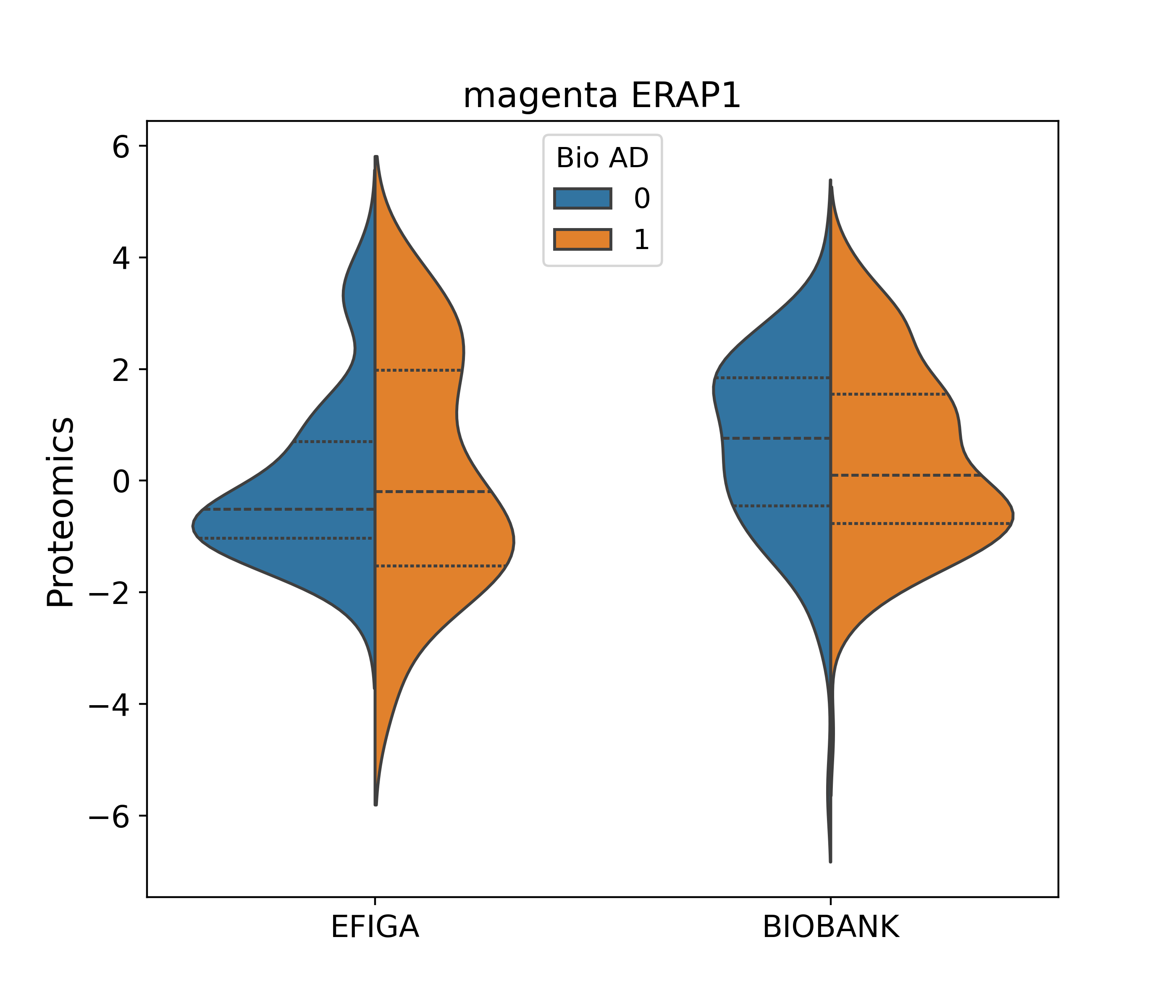

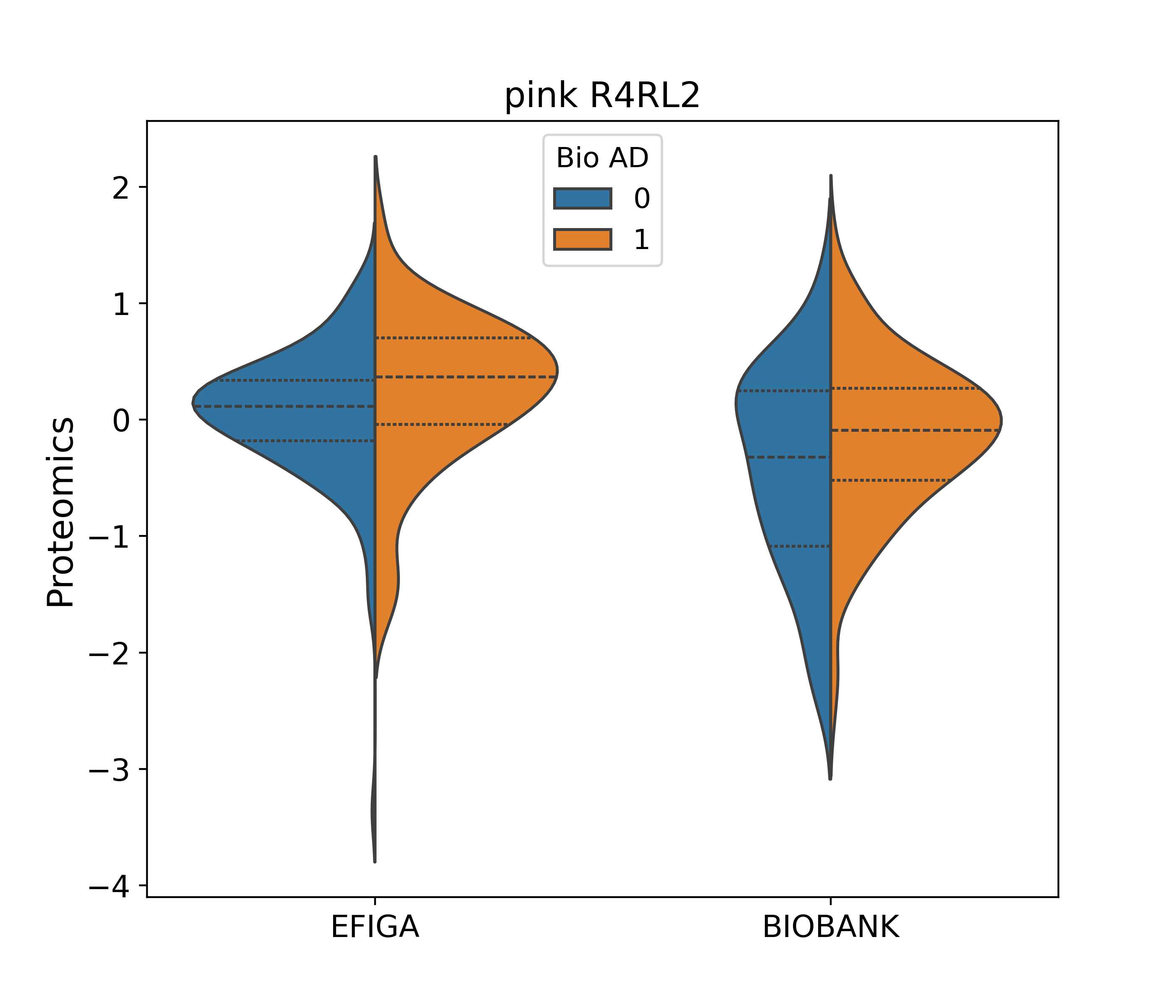

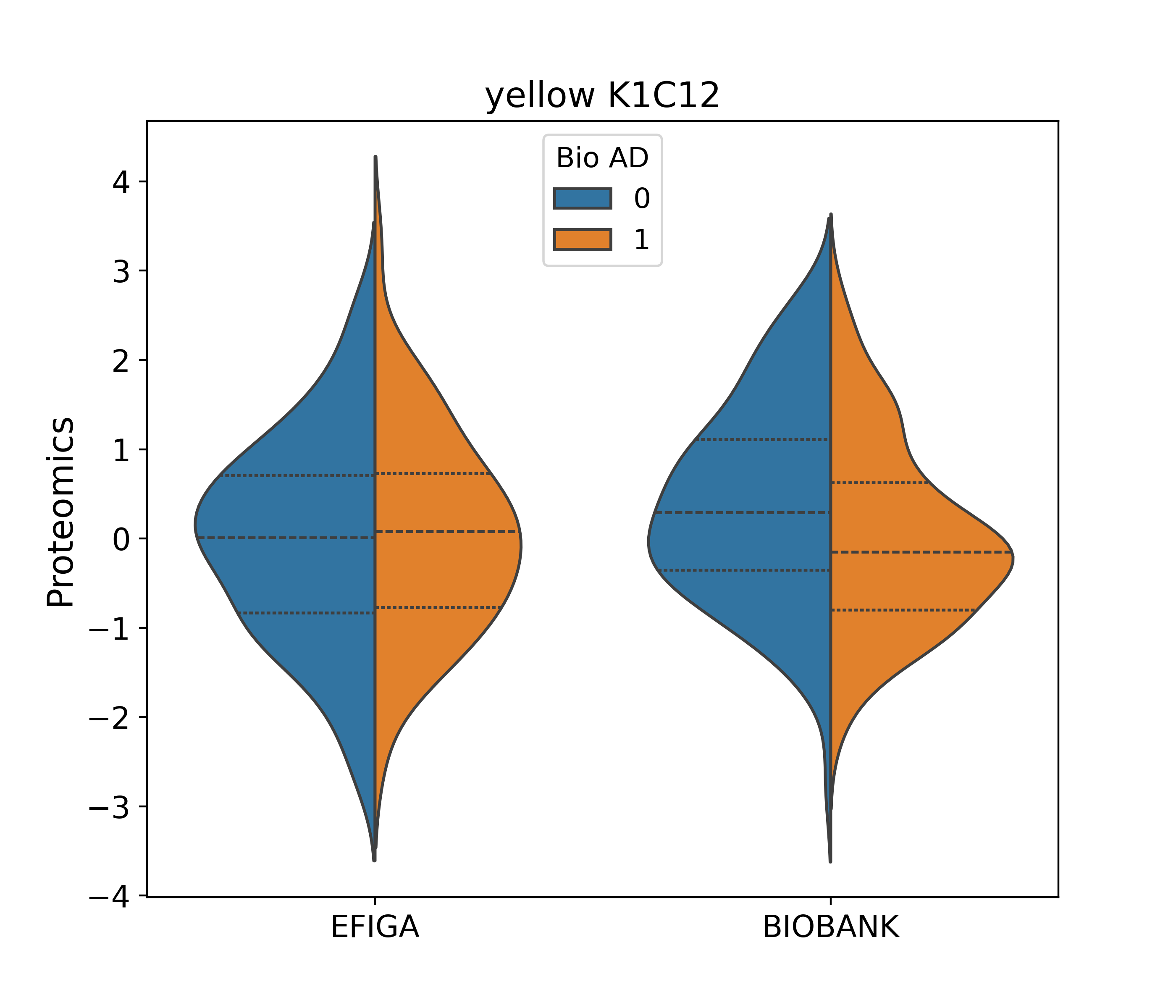

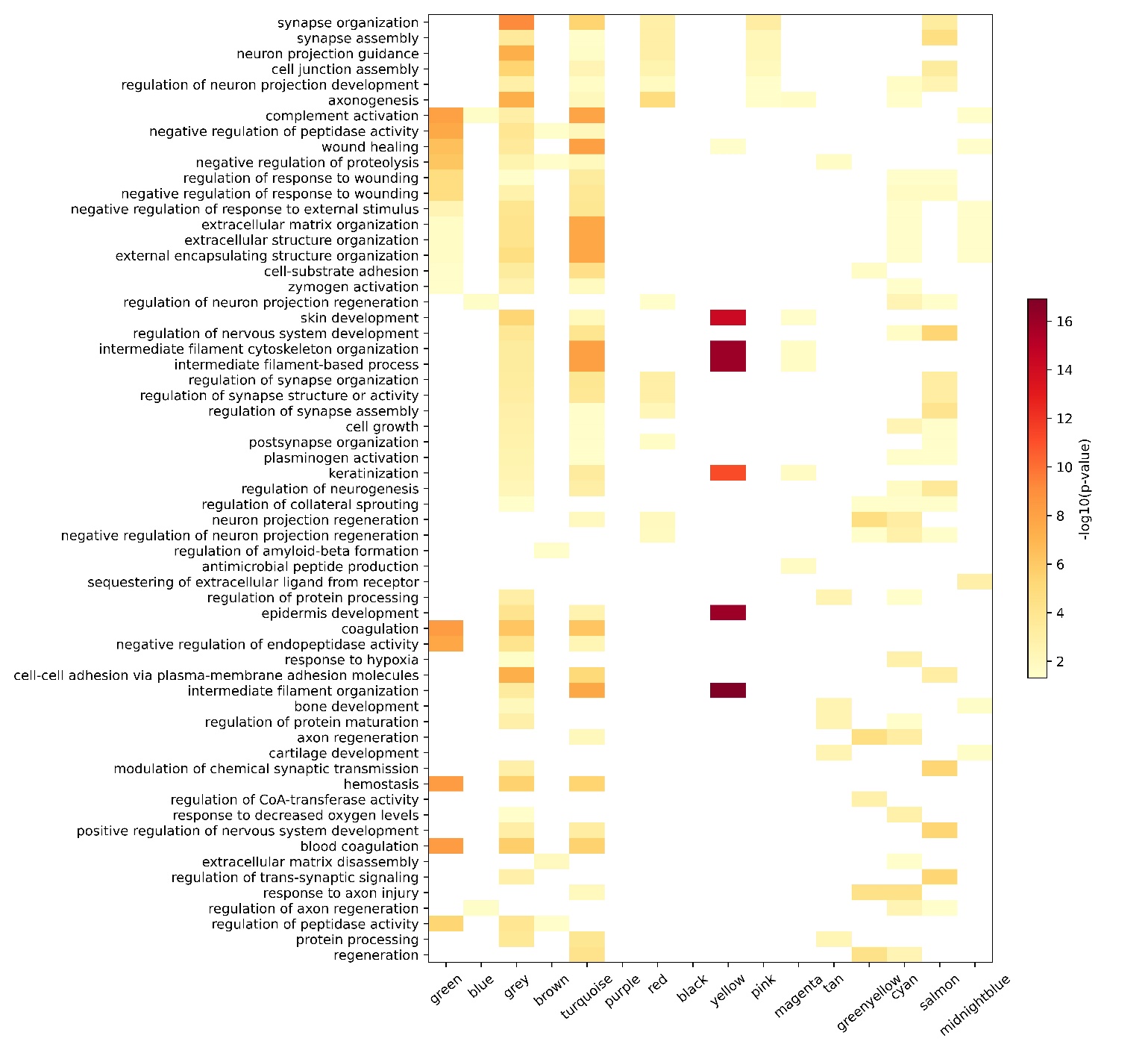
**Figure S4**

Pathways enriched within co-expression modules

**Figure S5**
Volcano plots of proteins associated with a) amyloid burden, b) tangle load, c) pathological AD diagnosis (using NIA-Reagan criterion) and d) global pathology measured in five different brain regions.

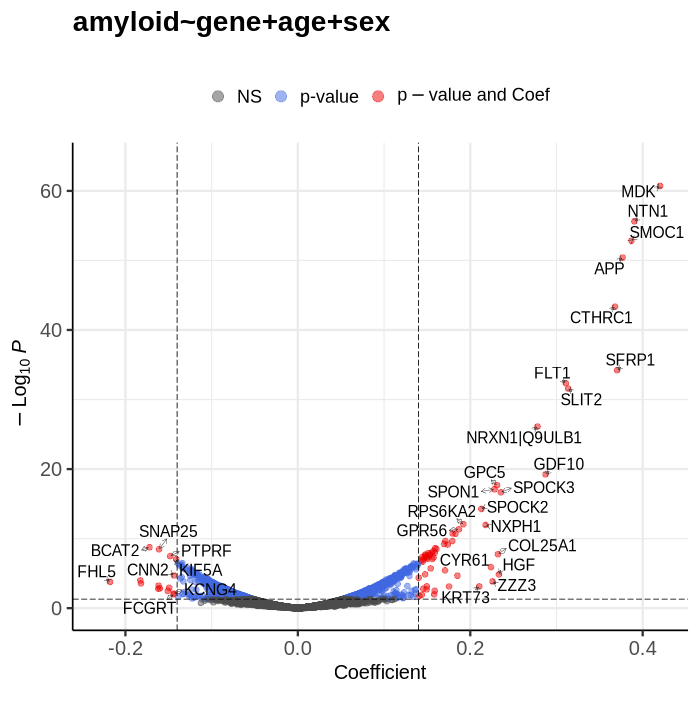

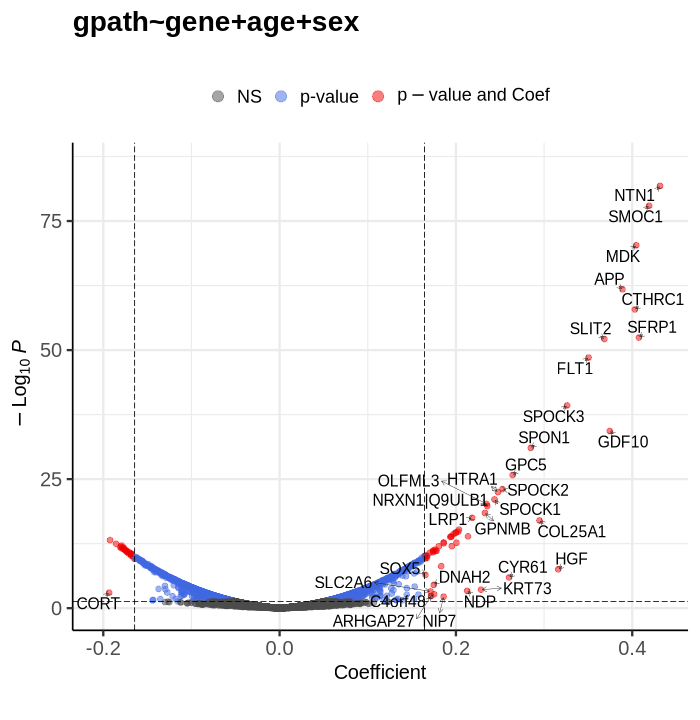

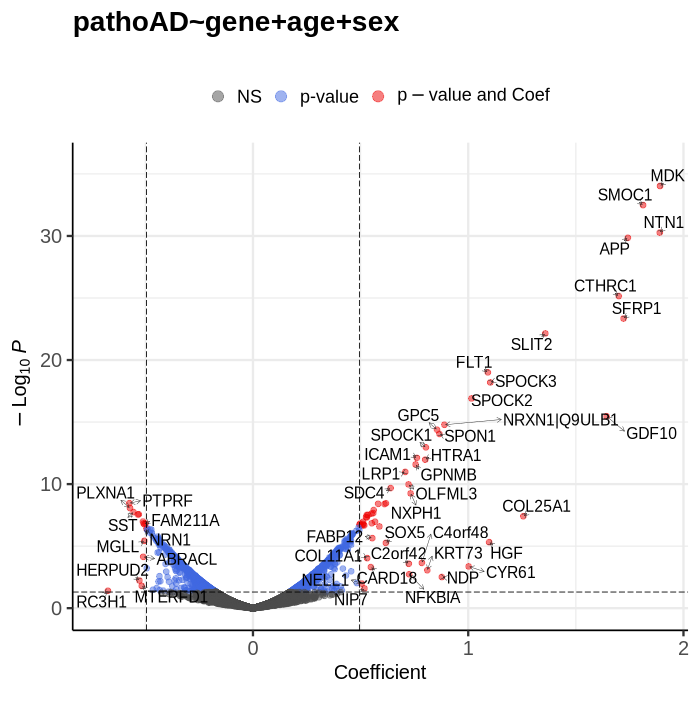

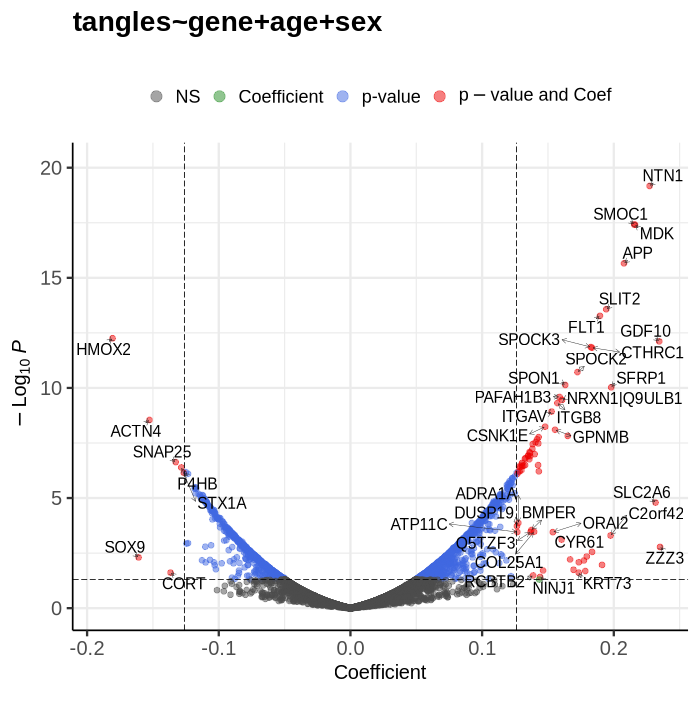

**Figure S6: Distribution of CSF biomarkers by clinical AD status in all cohorts.**

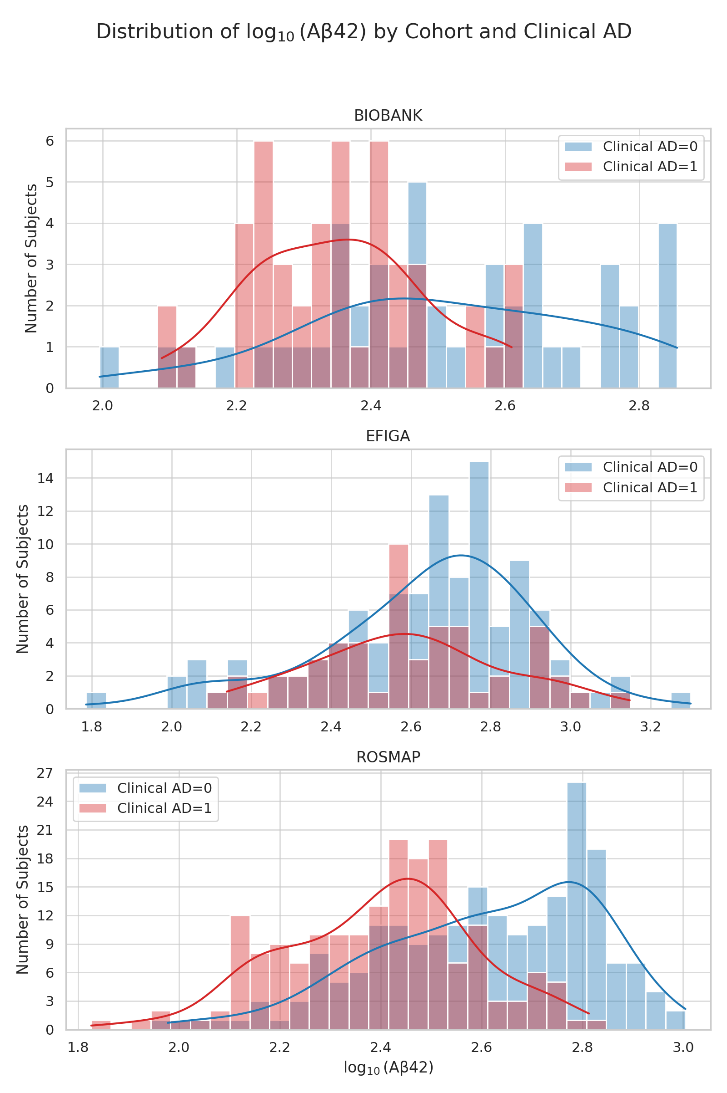

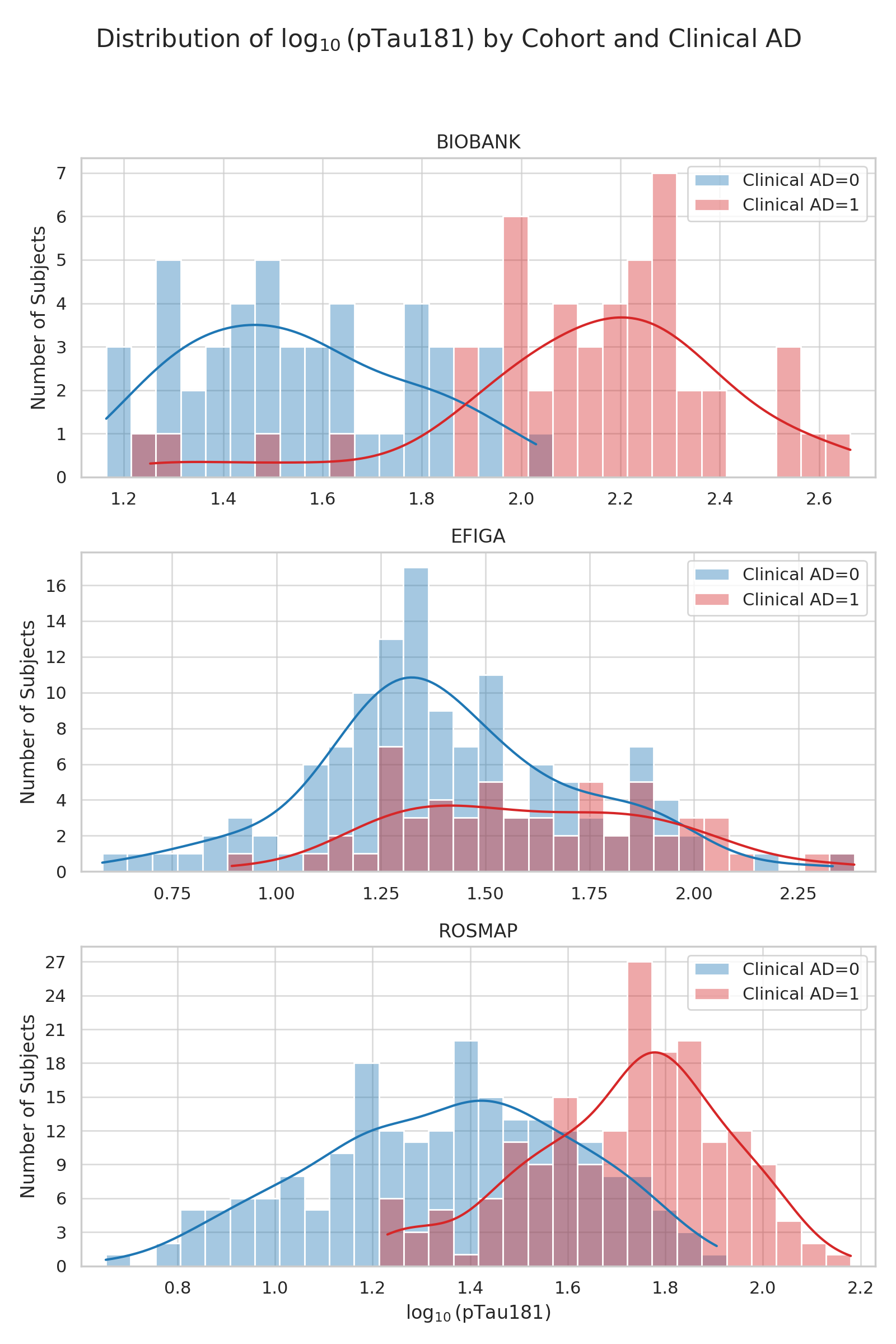

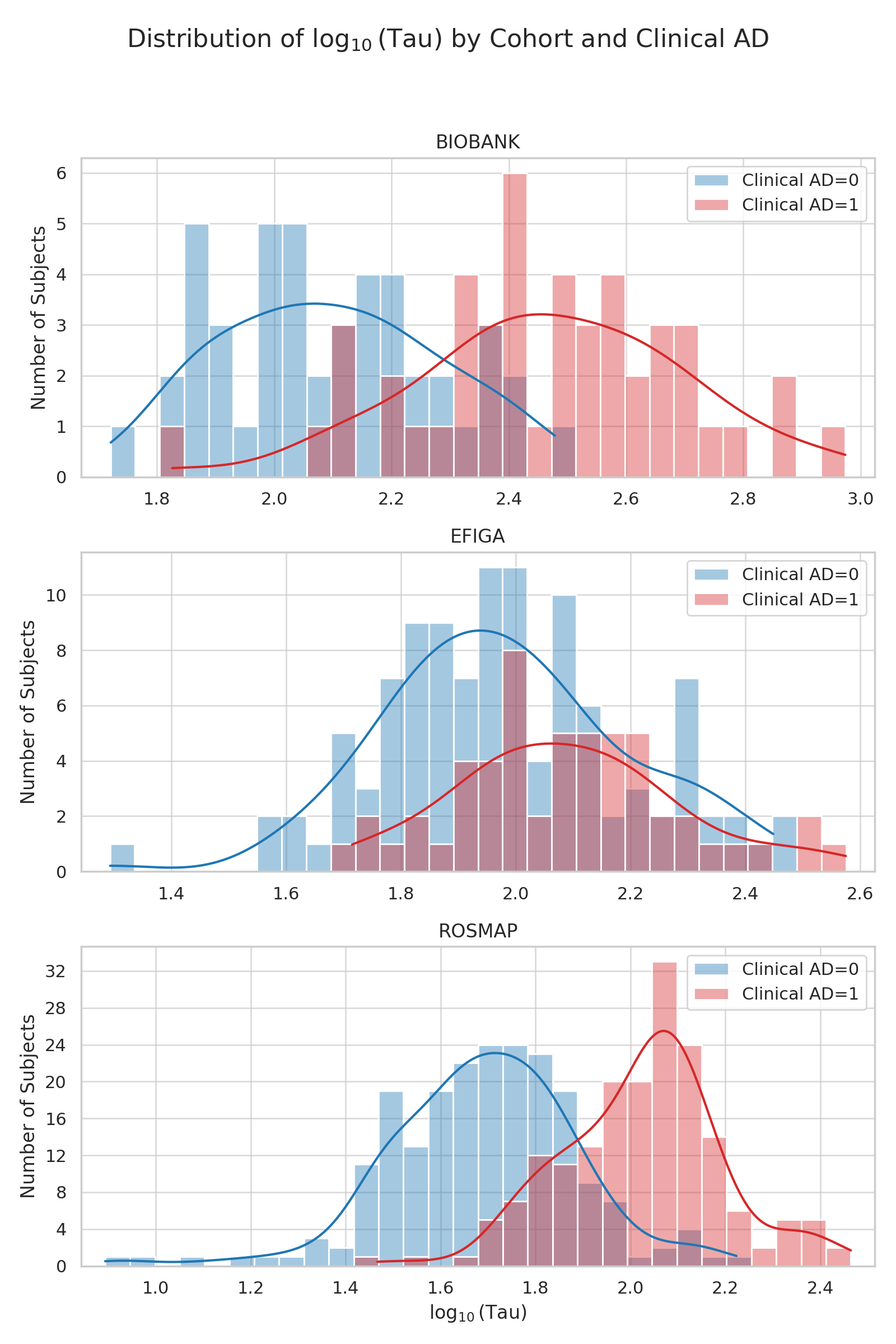

**Figure S7: Cell-type specific expression of ENPP2 and SPP1 (Osteopontin) in human and zebrafish datasets**

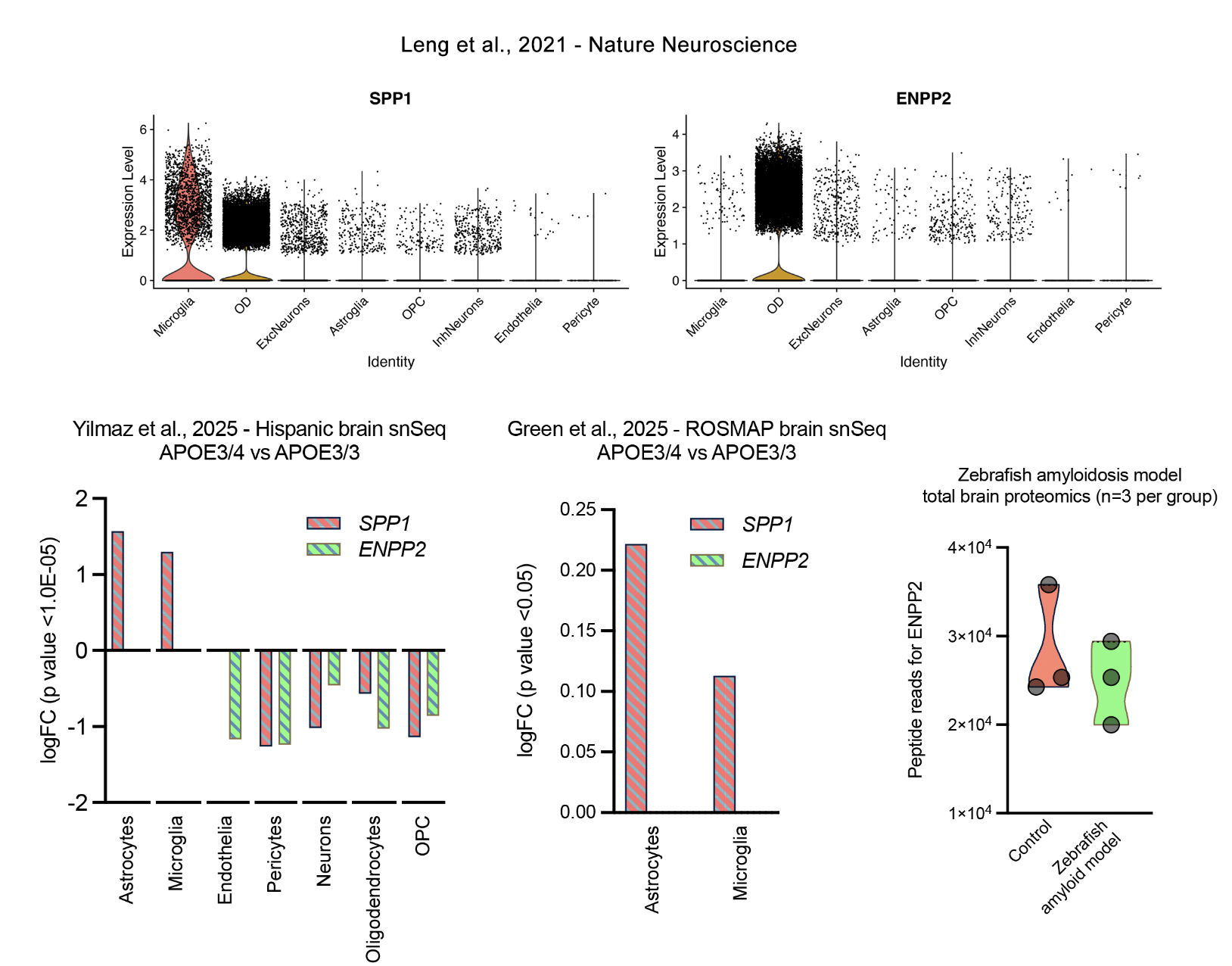
